# Poor immunogenicity upon SARS-CoV-2 mRNA vaccinations in autoimmune SLE patients is associated with pronounced EF-mediated responses and anti-BAFF/Belimumab treatment

**DOI:** 10.1101/2023.06.08.23291159

**Authors:** Caterina E. Faliti, Fabliha A. Anam, Narayanaiah Cheedarla, Matthew C. Woodruff, Sabeena Y. Usman, Martin C. Runnstrom, Trinh T.P. Van, Shuya Kyu, Hasan Ahmed, Andrea Morrison-Porter, Hannah Quehl, Natalie S. Haddad, Weirong Chen, Suneethamma Cheedarla, Andrew S. Neish, John D. Roback, Rustom Antia, Arezou Khosroshahi, F. Eun-Hyung Lee, Ignacio Sanz

## Abstract

Novel mRNA vaccines have resulted in a reduced number of SARS-CoV-2 infections and hospitalizations. Yet, there is a paucity of studies regarding their effectiveness on immunocompromised autoimmune subjects. In this study, we enrolled subjects naïve to SARS-CoV-2 infections from two cohorts of healthy donors (HD, n=56) and systemic lupus erythematosus (SLE, n=69). Serological assessments of their circulating antibodies revealed a significant reduction of potency and breadth of neutralization in the SLE group, only partially rescued by a 3^rd^ booster dose. Immunological memory responses in the SLE cohort were characterized by a reduced magnitude of spike-reactive B and T cell responses that were strongly associated with poor seroconversion.

Vaccinated SLE subjects were defined by a distinct expansion and persistence of a DN2 spike-reactive memory B cell pool and a contraction of spike-specific memory cTfh cells, contrasting with the sustained germinal center (GC)-driven activity mediated by mRNA vaccination in the healthy population. Among the SLE-associated factors that dampened the vaccine responses, treatment with the monoclonal antibody anti-BAFF/Belimumab (a lupus FDA-approved B cell targeting agent) profoundly affected the vaccine responsiveness by restricting the *de novo* B cell responses and promoting stronger extra-follicular (EF)-mediated responses that were associated with poor immunogenicity and impaired immunological memory.

In summary, this study interrogates antigen-specific responses and characterized the immune cell landscape associated with mRNA vaccination in SLE. The identification of factors associated with reduced vaccine efficacy illustrates the impact of SLE B cell biology on mRNA vaccine responses and provides guidance for the management of boosters and recall vaccinations in SLE patients according to their disease endotype and modality of treatment.

## Introduction

The latest 2019 outbreak with a novel coronavirus named SARS-CoV-2 and the starting of a deadly coronavirus disease 2019 (COVID-19) pandemic called for a unique situation where mRNA vaccines were trialed in a relatively large cohort of healthy donor volunteers and their approval in the USA was issued as emergency use approval (EUA) from the Food and Drug Administration (FDA)^1,2^. The distribution of monovalent mRNA vaccines and their boosters’ administrations according to a 3-dose injection schedule resulted in an effective reduction of severe COVID-19 infections and deaths^3^, as well as protection from the ancestral virus and its variants of concern^4,5^.

Vaccines’ efficacy in the healthy population has been assessed by investigating both humoral and cellular-associated responses. When administered to the general population, the mRNA vaccines induce prolonged germinal center (GC) activity^6,7^, generation of high-affinity neutralizing antibodies (nAb), and the establishment of immune memory^8-11^, as well as *de novo* B cell engagement upon booster^12,13^. While GC activity can be detected at least up to 6 months post-vaccination, the rapid waning of circulating and neutralizing antibodies^14,15^ suggests that the formation of long-lived plasma cells (LLPC) might not be vigorous^16^. Although still under investigation, several correlates of protection (CoP) against reinfections and/or severe disease manifestations have been proposed. Alongside the quantification of anti-Receptor-Binding-Domain (RBD) titers and virus-neutralizing antibodies (nAb), also persisting memory B cells and T cells are believed to play critical roles during infections and have been proposed as CoP^17^.

SARS-CoV-2 mRNA vaccines are safe and effective in immunocompetent subjects^1,2,18^, but the exclusion of immunocompromised subjects from the phase 3 clinical trials restricted our understanding of their efficacy in these vulnerable groups. Moreover, the immunocompromised state has been associated with prolonged SARS-CoV-2 shedding and reduced virus clearance during infections^19,20^, therefore urging the necessity for long-term effectiveness of vaccines administered to subjects at higher risk of infections, that could also behave as potential carriers of new variants.

Previous studies have reported reduced immunogenicity of the COVID-19 vaccines administered to subjects diagnosed with a wide spectrum of autoimmune disorders^21-23^, including lupus^24^. Underlying, pre-existing immune system defects and the concomitant treatment with a combination of immunomodulatory drugs can predispose those subjects to poor immunogenicity upon certain vaccinations^25,26^ and a higher risk and severity of bacterial and viral infections^27^. However, there is still a paucity of information about the mechanisms controlling mRNA vaccines’ immunogenicity and long-term efficacy in immunocompromised, autoimmune lupus subjects.

In this study, we investigated the efficacy of SARS-CoV-2 mRNA vaccinations in a vulnerable population of immunocompromised, autoimmune patients diagnosed with systemic lupus erythematosus (SLE). B cells play central roles in SLE through antibody-dependent and - independent functions^28^ with an important cross-talk with T cells^29^ often associated with aberrant functions^30-32^. We and others have shown that B cell regulation is profoundly disturbed in SLE through the expansion of activated effector B cells and plasma cells, a profile that segregates with more severe disease and may reflect a naïve-derived extrafollicular endotype distinct from a germinal-center/memory endotype^33,34^. Notably, severe COVID-19 infections induce prominent EF B cell differentiation and expansion of antibody-secreting cells (ASC) carrying autoreactive potential^35^, also characteristic of severe flaring SLE^36^.

Here, we performed in-depth analyses of both serological and cellular vaccine-mediated responses in all the participants of our study and correlated their immune endotypes, memory responses as well as lupus-associated treatments. We observed that, despite seroconverting, SLE patients had reduced titers and avidity of their neutralizing IgG and lacked coordinated activity between the memory T and B cell compartments. Subjects’ immune landscapes (immune endotypes) and antigen-specific EF B cells and T cell responses were strongly associated with negative or low vaccine responses, and a greater expansion of DN2 and DN3 B cells (a hallmark of EF activity) was observed in the SLE cohort, at both effector and late memory time points. Among other factors, the administration of Belimumab – the first SLE-specific FDA-approved monoclonal antibody targeting BAFF cytokine to reduce the survival of autoreactive B cells^37-39^– was associated with the poorest mRNA vaccine immunogenicity, even after a 3^rd^ booster dose. In all, our results shed light into mRNA vaccine efficacy in SLE; unravel the mechanisms underlying different types of responses; offer original insight into the relevance of disease B cell endotypes for vaccine responses; and provide guidance for strategies to optimize vaccine responses.

## Results

### Donors’ cohort characteristics and study design

To study the impact of preexisting autoimmunity and immunomodulatory therapy on SARS-CoV-2 vaccination outcomes, we enrolled donors that received two (Vax1+Vax2) or three (Vax3) doses of monovalent (Wuhan-Hu-1, WA.1) BioNTech/Pfizer or mRNA-1273/Moderna vaccines. The cohort consisted of both SLE patients (n=79 subjects, n=10 pre-pandemic, n=69 vaccinees) and age-and sex-matched healthy donor control groups (HD, n=64, n=8 pre-pandemic and n=56 vaccinees) collected between March 2021 and October 2022. Samples were mainly cross-sectional collections, with some longitudinal follow-ups as indicated in the diagrams (Table 1 and Extended Data Fig. 1). A total of 256 HD and 212 SLE sera or plasma samples were serologically evaluated for their vaccine-dependent antibody responses, and a total of 161 HD and 182 SLE paired PBMC samples were assessed for their antigen-specific B and T cell responses using high-dimensional flow cytometry combining antigen reactivity and deep immune profiling (Extended data Fig. 2).

### Lower seroconversion upon primary mRNA vaccination in SLE patients

To assess the level of seroconversion in our two cohorts and to define the specificity, kinetics, and persistence of circulating antibodies recognizing different portions of the mRNA-coded proteins, we performed an isotype-specific plasma screen^40^ against the following SARS-CoV-2 targets: S1 and S2 subunit domains of the spike protein, receptor-binding-domain (RBD) of the S1 subunit, as well as the N-terminal domain (NTD) (Extended Data Fig. 3a). Nucleocapsid (N) specific IgG were also tested to exclude prior infections (Extended Data Fig. 3b). Overall, IgM responses were minor contributors to the spike-reactivity in the HD cohort and increased in the SLE cohort (Extended Data Fig. 3c, d). IgA-specific responses were largely induced against the spike and RBD domains (Extended Data Fig. 3e, f) with highly similar responses between the groups. As previously reported for HD^41^, we observed in both cohorts a predominant IgG-mediated anti-spike, anti-RBD response (Extended Data Fig. 4, Fig. 1a-g).

**Fig. 1.**
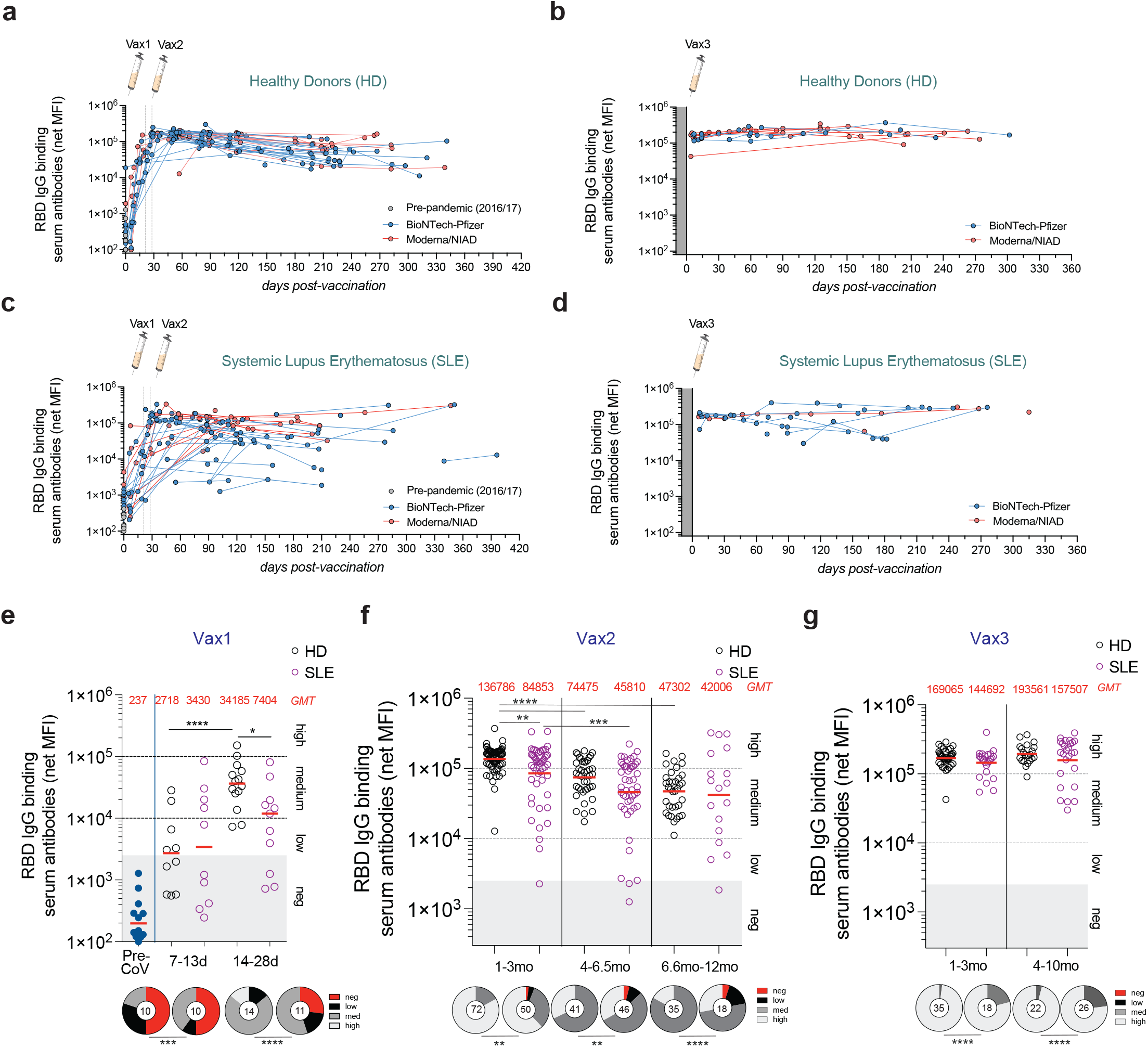
Serological evaluation of anti-spike vaccine-mediated antibodies responses. **a**-**d**, Luminex-based detection of RBD IgG binding serum antibodies (net MFI values) in the cohort of HD and SLE shown for each vaccine administration. Each dot represents a sample. Connecting lines show longitudinal collections. Comparisons between mRNA vaccines is shown for the BioNTech-Pfizer (aqua) and Moderna/NIAD (salmon) for Vax1+2 in the (**a**) HD and (**c**) SLE, and Vax3 for the (**b**) HD and (**d**) SLE. **e-g,** Clusters of IgG RBD titers based on binned time-points for samples collected on (**e**) Vax1, (**f**) Vax2 and (**g**) Vax3. Statistical analysis was performed with the Mann-Whitney U test and indicated when significant. Pie charts show the distribution of seronegative (MFIs 0-2500 MFI) and seropositive (low MFI 2500-10K, medium MFI 10k-100K, and high MFI >100k) values. The number of subjects is indicated in the pies, and the percentage of responders is compared with Chi-square.

Upon one mRNA vaccine dose administration (Vax1, 3-4 weeks), ∼85% of HD seroconverted, while the SLE patients had a lower seroconversion rate of ∼58%, with significantly more negative/low responders (Fig. 1e and Extended Data 4 a, b). Completion of the primary series of vaccines (Vax2) increased the seroconversion rate of the SLE (88%) and HD (100%), although reduced mean titers and overall increased number of negative/low responders remained in the SLE vaccinees (Fig. 1f and Extended Data 4 a, b). A booster dose (Vax3) normalized the mean titers of IgG RBD between the two cohorts (Fig. 1g and Extended Data 4 a, b).

### Reduced IgG competitiveness and neutralization potency and breadth in the SLE cohort

The functional activity of the circulating IgG was determined by a competitive ELISA assay to assess their efficiency in blocking the interaction of rec-hRBD with its receptor rec-hACE2 (Fig. 2a, b)^42^. Interestingly, while overall anti-RBD titers were only mildly reduced in SLE patients (Fig. 1), the SLE group displayed significant impairment in the ability to block ACE2 binding across most time points (Fig. 2b). Despite medium/high RBD IgG titers, a greater frequency of SLE samples was enriched in either non-competitive or low competitors IgG (Fig. 2c, d) which are usually low avidity IgG^42,43^ that might arise from ineffective affinity maturation processes.

**Fig. 2.**
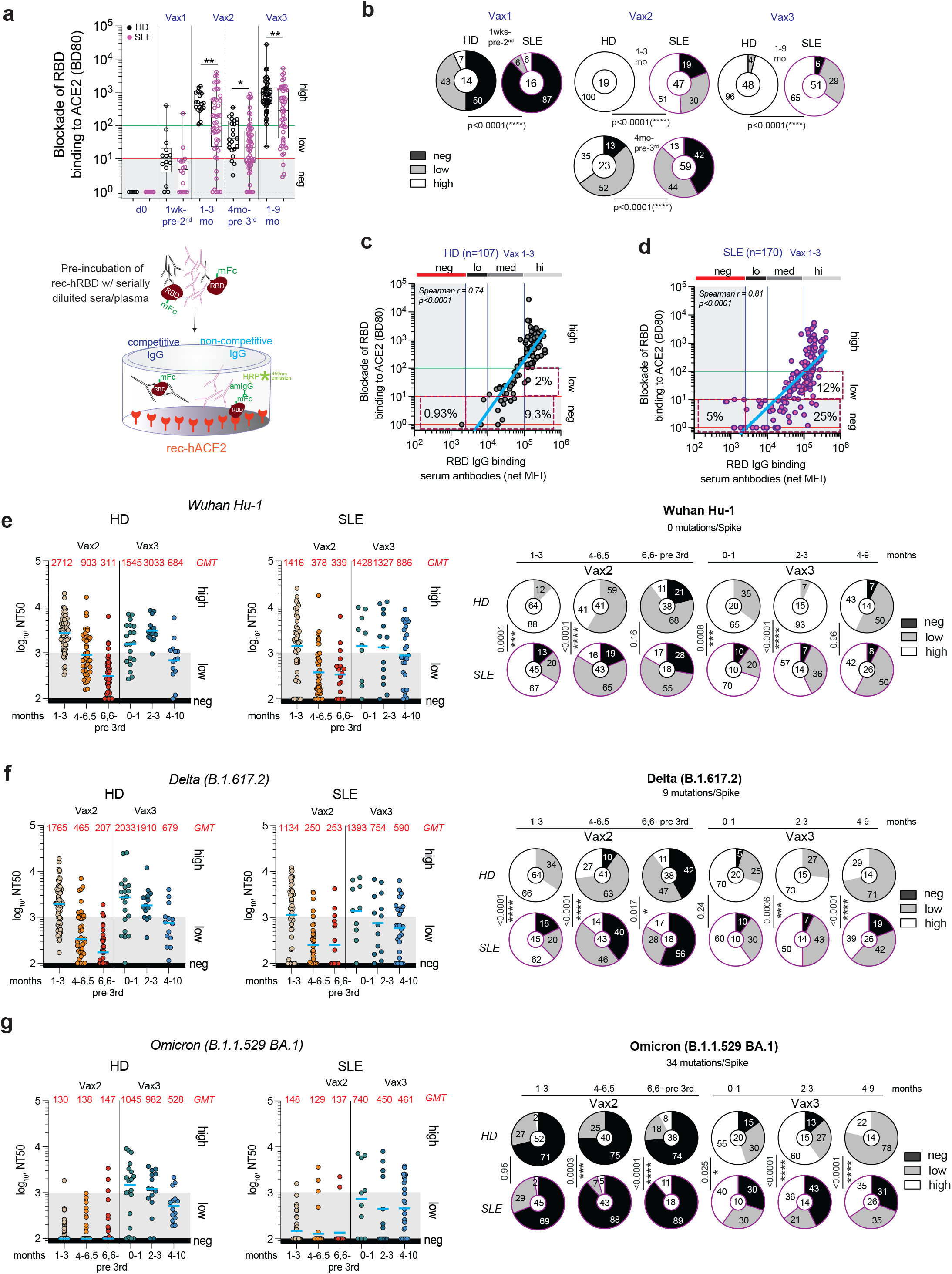
Reduced neutralization and breadth in the cohort of vaccinated SLE. **a**, ELISA determination of Ab-mediated inhibition of SARS-CoV-2 RBD binding to solid phase ACE2. The graph shows the reciprocal plasma or serum dilution that blocks RBD’s 80% binding (BD80) to human ACE2. BD80 log values are shown as negative (0-1), low (1-2), and high (>2). Each dot represents a tested sample. Statistical analysis was performed using the Mann-Whitney U test. **b**, Pie graphs showing the frequency and statistical comparison of competitive IgG in the two cohorts. Chi-square with Fisher’s test. The number in the circles indicates the total number of samples tested, while the numbers in the pies show the relative percentage of the negative (black), low (grey), and high (white) values. **c**, **d**, Graphs showing the linear correlation between the values of the blocking of RBD binding to ACE2 (BD80) and total RBD IgG binding antibodies in the same sample tested from (**c**) HD and (**d**) SLE vaccinated subjects. **e**, **f, g,** Pseudo-viral neutralization *in vitro* assay performed with plasma samples isolated from vaccinated subjects after vaccines 2 and 3. The graphs show the neutralizing titers inhibiting 50% of the viral growth (NT50) tested for the SARS-CoV-2 (**e**) Wuhan-Hu-1 wild-type, (**f**) Delta (B.1.617.2) and (**g**) Omicron (B.1.1.529 BA.1) strains. Each dot in the bar plots represents an individual sample tested. Pie charts comparison of negative and low and high neutralizers. Statistical comparison was performed using the Chi-square with Fisher’s test. The number in the circles indicates the total number of samples tested, while the numbers in the pies show the relative percentage of the negative (black), low (grey), and high (white) values.

To corroborate the ELISA results, we performed pseudoviral neutralization assays^44^ against SARS-CoV-2 Wuhan-Hu-1 WT (WA.1), Delta (B.1.617.2), and Omicron (B.1.1.529 BA.1) strains. As predicted by the blockade assay, SLE patients displayed impaired neutralization activity against those viruses relative to their HD counterparts, with significantly lower neutralization titers and reduced breadth in samples collected in the months following both the primary series and boost (Fig. 2e-g).

Interestingly, neutralizing titers decayed significantly faster in HD relative to SLE, with significant differences for Wuhan-Hu-1 WT (half-life of 38 days for HD vs 73 days for SLE; p=0.03), and Delta (half-life of 39 vs 68 days; p=0.03), after Vax3^45^ (day 42 post-vaccination and onwards). These differences were not detected for these strains during the shorter follow-up interval after Vax2 nor for Omicron strain after Vax3.

While the RBD domain of the S1 has been described as a main target of broadly neutralizing antibodies (bnAb), other non-RBD structural proteins of the spike, namely S2 and NTD, can harbor neutralizing epitopes. The S2 domain, harboring more conserved epitopes and some level of cross-reactivity with other coronaviruses, was targeted similarly by HD and SLE IgG (Extended Data Fig. 4a-c), while the IgG reactivity towards the NTD domain was lower in the SLE cohort, potentially compromising the control of viral escape^46,47^ (Extended Data Fig. 4c, d).

Overall, the evaluation of serological responses to mRNA vaccination revealed a defective primary response in SLE that requires vaccine boosters for full seroconversion. Moreover, SLE responses were qualitatively defective in and displayed lower breadth of neutralization.

### Defective priming, and distinct B cell responses to mRNA vaccination in SLE

To evaluate the magnitude of cellular responses and the identity of the effector/memory B cells induced upon mRNA vaccination, we performed comprehensive immunophenotypic profiling with a high-dimensional 28-color flow panel that provided tetramer-based detection of WA.1 spike and RBD-reactive B cells (Extended Data Fig. 2a and Fig. 3a, b). During the post-Vax1 priming phase, 62% of HD subjects mounted an early anti-spike B cell response (Fig. 3c), with ∼35% of those also binding to the RBD tetramer (Fig. 3d). Virtually all the HD subjects displayed a higher and persistent anti-spike, anti-RBD recall response to Vax2 and Vax3 (Fig. 3c, d). The SLE group had a significantly lower proportion of responders after priming and within memory recall responses, with ∼10-30% of patients failing to generate a detectable B cell response at any time (Fig. 3c, d).

**Fig. 3.**
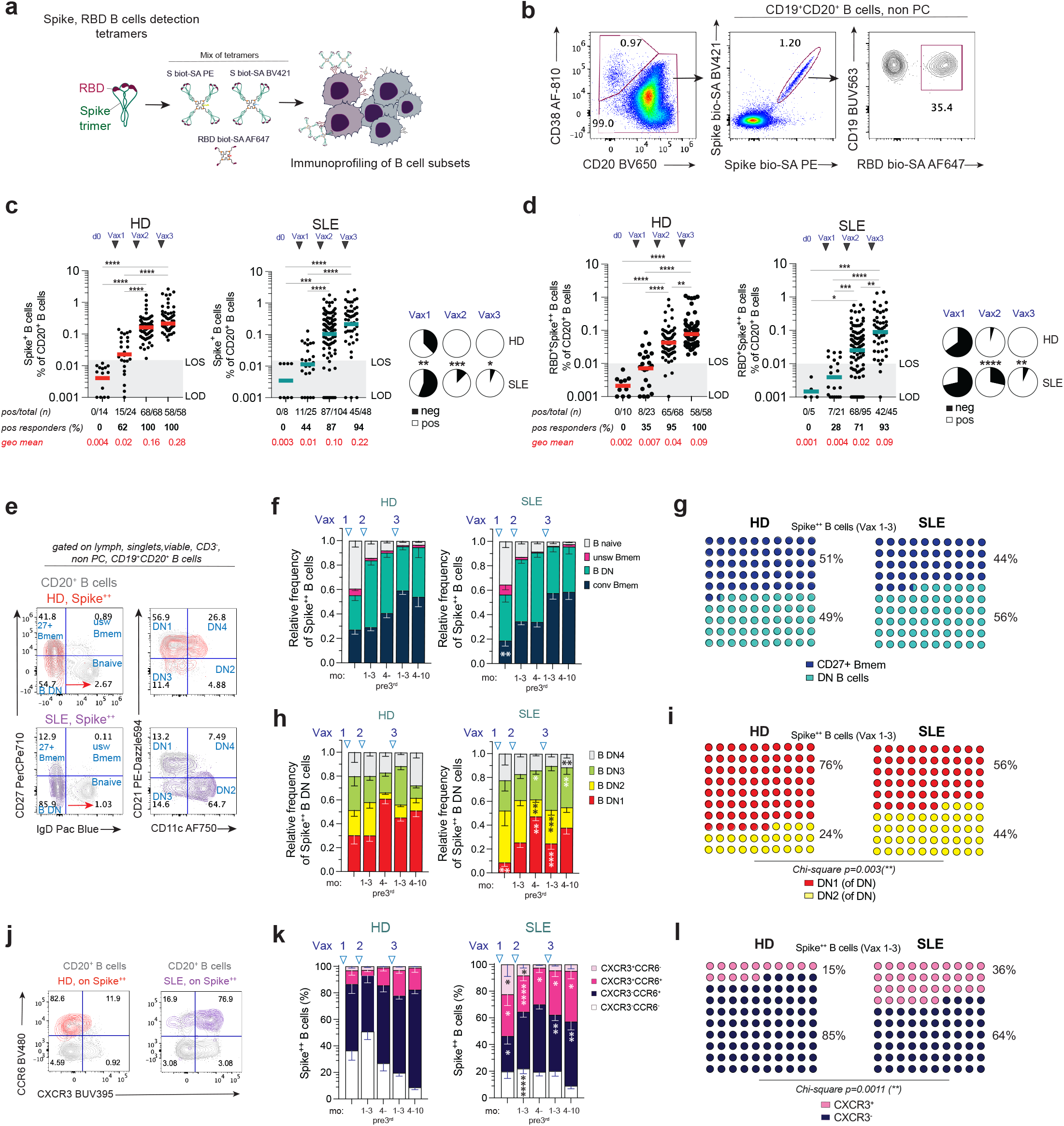
Lower magnitude of antigen-specific memory B cells and greater DN2 expansion in the vaccinated SLE cohort. **a**, Cartoon showing the *ex-vivo* tetramer-based detection of spike, RBD reactive B cells and high dimensional flow immunoprofiling of B cells from PBMCs **b,** Representative FACS plots showing the gating strategy applied to characterize the total CD19^+^CD20^+^ B cells (excluding the CD20^-^ CD38^hi^ plasma cells) binding to dual-tetrameric spike probes and tetrameric RBD probe. **c, d,** Quantification of total (**c**) spike-specific and (**d**) RBD-specific B cells shown as the frequency of CD20^+^ B cells in the HD and SLE cohorts. Each dot represents an individual sample tested at baseline (d0) and after receiving one (Vax 1), two (Vax 2), or three (Vax 3) vaccine doses. Pie charts comparisons and Chi-square with Fisher’s test are shown. **e,** Representative FACS plots showing the characterization of spike-reactive CD20^+^ B cells subsets based on the expression of IgD and CD27 to define B naïve (IgD^+^CD27^-^), unswitched B memory (IgD^+^CD27^+^), conventional B memory (IgD^-^CD27^+^) and double-negative B cells (DN, IgD^-^CD27^-^) cells. CD21 and CD11c markers are used to further define the DN subsets as DN1 (CD21^+^CD11c^-^), DN2 (CD21^-^CD11c^+^), DN3 (CD21^-^CD11c^-^) and DN4 (CD21^+^CD11c^+^) cells. **f,** Bar graphs showing the relative frequency of spike^++^ B cell subsets of IgD and CD27 in HD and SLE vaccinees. Mann-Whitney U test the statistical significance of SLE B cell subsets populations compared to HD frequencies is shown in the SLE graphs for each specific group of the column, at the indicated Vax time point. **g,** 10×10 dot plots showing the comparison of conventional CD27^+^ (midnight blue) and DN (teal green) spike^++^ B cells, and the relative frequencies are shown for all vaccinated samples combined from HD and SLE cohorts. **h,** Bar graphs showing the relative frequency of spike^++^ B cell subsets of DN B cell subsets in HD and SLE vaccinees. Mann-Whitney U test of the statistical significance of SLE B cell subsets populations compared to HD frequencies is shown in the SLE graphs. **i,** 10×10 dot plots showing the comparison of conventional DN1 (red) and DN2 (yellow) spike^++^ B cells, and the relative frequencies are shown for all vaccinated samples combined from HD and SLE cohorts. Chi-square with Fisher’s test. **j**, Dot plots representative of chemokine receptors CCR6 and CXCR3 expression of total and spike^++^ B cells. **k**, Bar graphs showing the distribution of CCR6 and CXCR3 expressing spike-reactive B cells of the HD and SLE cohort. Mann-Whitney U test the statistical significance of SLE B cell subsets populations compared to HD frequencies is shown in the SLE graphs. **l,** 10×10 dot plots showing the comparison of the total CXCR3^+^ (light pink) and CXCR3^-^ (dark blue) spike^++^ B cells, and the relative frequencies are shown for all vaccinated samples combined from HD and SLE cohorts. Chi-square with Fisher’s test. When indicated, LOD=limit of detection set to logarithmic 0.001 for B cells and 0.003 for T cells. LOS=limit of sensitivity, based on median values of baseline + 2 x SD.

B cell lymphopenia is common in SLE owing to both, disease activity and therapy. To account for this variable, we compared ag-specific frequencies of total CD19^+^ B cells between HD and SLE (Extended Data Fig. 6a). Both total and ag-specific circulating B cells were reduced in SLE, with enrichment of patients with extremely low B cell frequency (<1%) and low-spike^+^ cells (<0.0022%) (Extended Data Fig. 6b). Normalization of ag-specific reactivity in this way confirmed that the SLE cohort carried more negative responders that the HD group (Extended Data Fig. 6c, d). Contrary to the greater durability of their antibody response, the frequency of spike-reactive B cells calculated on viable lymphocytes declined more rapidly in SLE after Vax2 (half-life (95% CI), calculated decay p=0.009). However, no significant differences between the groups were observed after Vax3.

### Novel CD27^-^ memory populations contribute disproportionately to spike memory B cells in SLE

In addition to the magnitude and durability of the antibody response, an optimized vaccine should also induce robust effector and memory B cells to provide both short-term and long-term protection, respectively. Despite the growing complexity of these compartments, thus far, only plasmablasts and CD27^+^ memory responses have been largely investigated. Since different effector and memory pathways may be induced in different types of SLE^35,48^, we sought to further interrogate the expanded pool of antigen-reactive B cells and define their dynamics. Our cytometry approach measured the distribution of spike-reactive B cells among the larger parental populations defined by the expression of IgD and CD27: naïve, CD27^+^ unswitched memory and CD27^+^ isotype switched memory as well as IgD/CD27 double-negative cells (DN). DN B cells were further fractioned into four specific subsets (DN1-4) determined by the expression of CD21 and CD11c (full gating strategy Extended Data Fig. 2a, Fig. 3e).

CD20^+^ spike^+^ B cell responses to Vax1 included in both cohorts similarly large fractions of naïve B cells (∼40%), that rapidly contracted over subsequent vaccinations and time points (Fig. 3f). The IgD^+^ unswitched memory cells also contributed to a small portion of the total antigen-induced population that rapidly contracted (Fig. 3f). The initial priming responses also included significant fractions of early CD27^+^ switched memory cells and DN cells, the latter representing naïve-derived effector responses as we had previously reported for autoreactive B cells in SLE and severe COVID-19 infections^36,48,49^. Notably, DN cells dominated the spike response early in SLE and remained dominant in both groups prior to booster vaccination, presumably reflecting memory cells induced by Vax1 and effector activated memory cells induced by Vax2^50,51^. In both groups, conventional CD27^+^ memory cells dominated the response post-booster but with persistence of substantial fraction of DN memory cells (Fig. 3f, g).

Assessment of the DN subsets distribution (DN1-4) is an indicator of origin and function. Specifically, we have previously associated DN1 B cells with conventional CD27^+^ memory pathways^52^, and shown that they represent a large majority of DN cells in HD. In contrast, DN1 become a much lower fraction in acute SLE and severe COVID-19 infection, where DN2 and DN3 B cells become dominant. In these acute situations, DN2 and DN3 cells are considered EF naïve-derived effector cells^35,36,49,53,54^. However, little is known about the contribution of DN subsets to effector and memory vaccination responses and specifically, to mRNA vaccination. In this study, DN1 strongly dominated spike-specific response in HD across all time points. In contrast, the SLE response was dominated by DN2 cells in the early response to Vax1 and Vax 2 and remained significantly higher in SLE relative to HD in post-Vax 3 (Fig. 3h, i). DN3 cells also contributed to different phases of the response and were significantly expanded at late time points post-Vax2 and Vax3 in SLE (Fig. 3h).

We next analyzed Ig-surface expression in memory cells (Extended Data Fig. 7a). As predicted by the serology, IgG responses dominated the spike-reactivity (Extended Data Fig. 7 b-e), while IgM and IgA memory B cells represented a smaller portion of the response (Extended Data Fig. 7 b-d). Interestingly, IgM^+^ memory B cells were increased particularly in early post-Vax2 and Vax3 phases, accounting in several patients for more than over 20% of all antigen-reactive CD20^+^ B cells (Extended Data Fig. 7c). Additionally, SLE spike reactivity was associated with a smaller and delayed expansion of IgG^+^ B cells upon recall doses (Extended Data Fig. 7 b, e).

Spike-reactive B cells were also qualitatively different between the two cohorts in terms of their expression of CCR6 and CXCR3 chemokine receptors (Fig. 3j-l). In HD, we observed a predominance of CCR6^+^ spike B cells, that expanded within subsequent vaccinations (Fig. 3k, l). In contrast, SLE spike B cells were highly enriched in CXCR3^+^ cells (Fig. 3k, l).

### Impaired activation and persistence of vaccine-induced T cells in SLE

We also sought to define the magnitude and kinetics and differentiation of vaccine-induced CD8^+^ and CD4^+^ T cells upon mRNA vaccination. To this end, we used an *in vitro* system - suitable to score the frequency of antigen-reactive T cells - that relies on the incubation of cells with mega-pools (MPs) of peptides and allows their quantification via their expression of *in vitro* Activation-Induced Markers (“AIM” assay)^9,55-57^ (Fig. 4a and Extended Data Fig. 8 a-b). Cells were stimulated with the pool of spike overlapping peptides spanning the entire protein (WA.1, spike aa 5-1273), or with peptides from the HA H1N1 (A/California/04/2009), an unrelated control for viral T cell reactivity. Twenty-four hours after the incubation, samples were analyzed to score the magnitude of antigen-reactive AIM^+^ cells, using a combination of the expression of CD69 and 41BB for CD8s and CD40L and OX40 for CD4s. T helper CD4s were classified based on their expression of chemokine receptors and their polarization was further investigated in both the CXCR5 negative or positive (cTfh) compartments (Fig. 4b and Extended Data Fig. 8b).

**Fig. 4.**
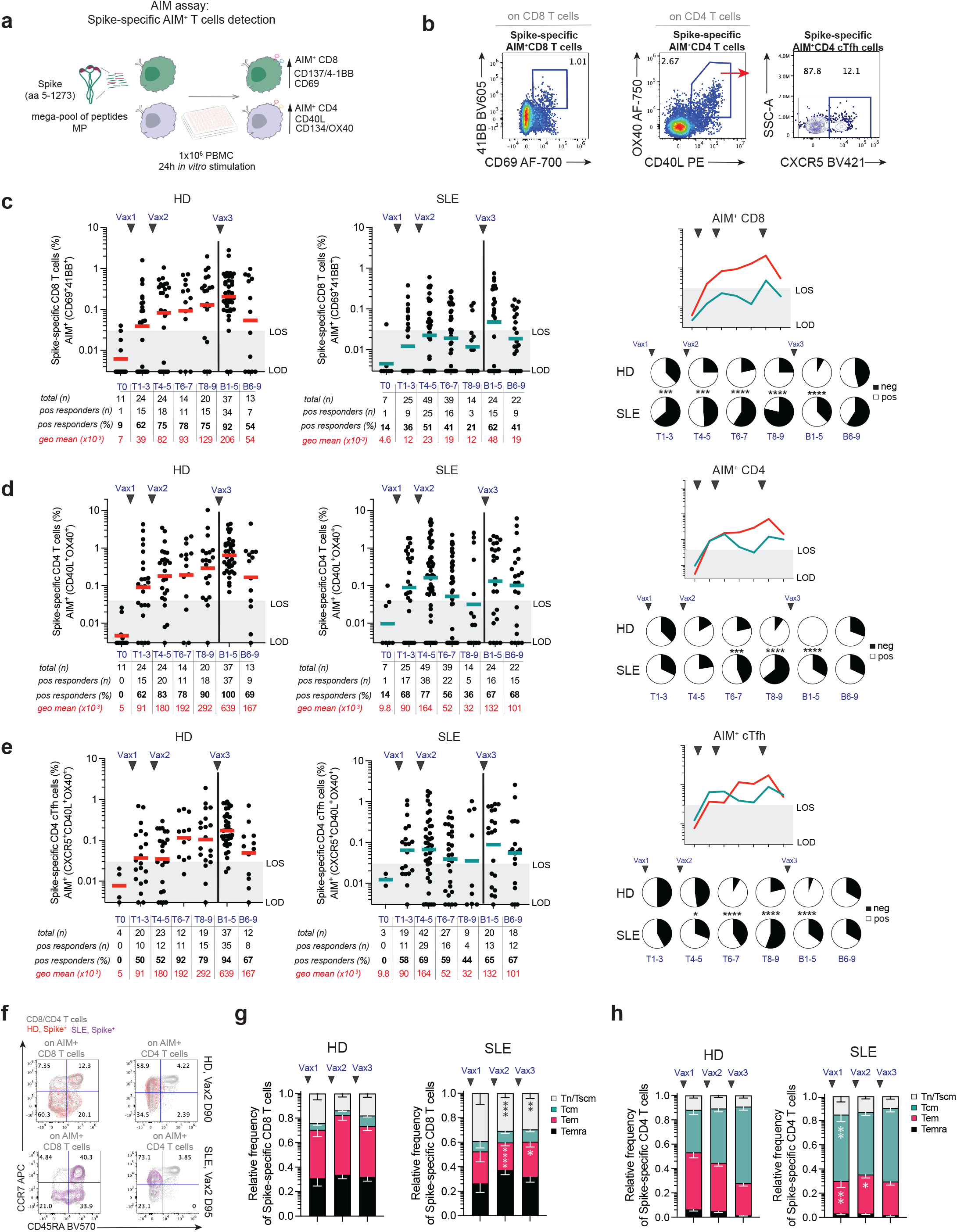
Lower T cell reactivity in SLE receiving the SARS-CoV-2 mRNA vaccines. **a**, Cartoon showing the 24h AIM assay-based detection of antigen reactive T cells upon PBMC incubation with a mega-pool of spike-derived peptides and flow cytometry analysis of surface-expressed markers of activation and immune profiling. **b,** Representative FACS plots showing the gating strategy applied to characterize the AIM^+^ spike reactive CD8 (41BB^+^CD69^+^) or AIM^+^ spike reactive CD4 (OX40^+^CD40L^+^) and AIM^+^ spike reactive cTfh (CXCR5^+^ of AIM^+^CD4) among the CD3^+^ T cells. Scatter plots showing the frequency of spike-specific AIM^+^ T cells quantified at each indicated time point in the cohort of HD and SLE subjects for **c,** AIM^+^ CD8, **d,** AIM^+^ CD4, and **e,** AIM^+^ CD4 cTfh. Overlay graphs for the geometric mean values of the groups are shown within the pie charts representing the proportion of negative and positive responders. The Chi-Square test was performed to define significance. **f,** Representative dot plots showing the differentiation of AIM^+^ T cells using CD45RA and CCR7 expression. **g**, Bar plots showing the distribution of the four T cell subsets (Tn/Tscm, Tcm, Tem and Temra) of AIM^+^ CD8. **h**, Bar plots showing the distribution of the four T cell subsets (Tn/Tscm, Tcm, Tem and Temra) of AIM^+^ CD4. Statistical Mann-Whitney U test was applied to compare each subset of T cells between SLE and HD, and significance showed in the SLE bars. When indicated, LOD=limit of detection set to logarithmic 0.001 for B cells and 0.003 for T cells. LOS=limit of sensitivity, based on median values of baseline + 2 x SD.

In keeping with the B cell responses, lower T cell responses were observed in the SLE cohort, with impaired priming of AIM^+^ CD8 (Fig. 4c) and lower levels of both AIM^+^ CD8 and CD4 at memory and recall phases (Fig. 4c-e). The overall fold reduction in the SLE group compared to HD was 6.71 (AIM^+^ CD8s) and 3.15 (AIM^+^ CD4s) at Vax2 and 3.85 (AIM^+^ CD8s) and 2.92 (AIM^+^ CD4s) at Vax3. Furthermore, while broad AIM^+^ T helper cells polarization was similar between the two cohorts (Extended Data Fig. 8c, d), we detected an initial skewing of the primed SLE AIM^+^ cTfh into CCR6^+^ Tfh17 cells (Extended Data Fig. 8d, e), with a general reduction of the magnitude of the SLE AIM^+^ cTfh pool upon memory reactivated responses (Fig. 4e). We further defined populations of naïve and effector/memory T cells based on the expression of CD45RA and CCR7 (Fig. 4f and Extended Data Fig. 8b) and observed a general reduction in the pool of effector AIM^+^ SLE T cells (Fig. 4g, h).

Defective generation of HA-reactive memory T cells was not observed in the same SLE subjects (Extended Data Fig. 8f), although the AIM^+^ HA CD8s pool still failed to generate normal EM cells, suggesting the CD8 defects might be mediated by associated lupus-disease defects (Extended Data Fig. 8g)^58^.

### Poor vaccine-mediated responses in SLE are associated with a strong EF immune signature

To investigate the influence of B cell endotypes on different vaccine responses across cohorts, we used the mean values of the total RBD reactive IgG to classify all vaccinees enrolled in this study into 3 groups of responders: negative/low (V_NL_), medium (V_M_), or high (V_H_) vaccine responders (Fig. 5a). As expected, RBD IgG titers directly correlated with the potency of the WA.1 neutralization (Fig. 5b). SLE vaccinees that received 1 or 2 doses of vaccine were enriched in negative/low responders, and their neutralizing IgG potency upon Vax2 was still significantly lower than the HD. While the booster dose improved the responses in the SLE, only 78% reached high antibody titers relative to 97% of HD (Fig. 5 a, b). We then tested the hypothesis that in our SLE cohort, a predisposition to strong EF immune responses might be responsible for lower immunogenicity of mRNA vaccination. To address this question, we assessed patients for circulating cellular surrogates of EF activity identified in previous studies of SLE and acute COVID-19^36,48,49^. Based on these studies, we created an EF activity score including low DN1 frequency within the DN compartment, low cTfh signature, and expansion of DN2 B cells and plasma cells (Fig. 5c, Extended Data Fig. 9e). We observed that strong EF signatures were indeed associated with V_NL_ groups and were strongly over-represented in the SLE cohort where they negatively correlated with the vaccine responsiveness (Fig. 5d). Factors contributing to vaccine responses such as age, sex, or race did not appear to drive strong bias in the mRNA immunogenicity and vaccine responsiveness in both cohorts (Extended Data Fig. 9 a-d).

**Fig. 5.**
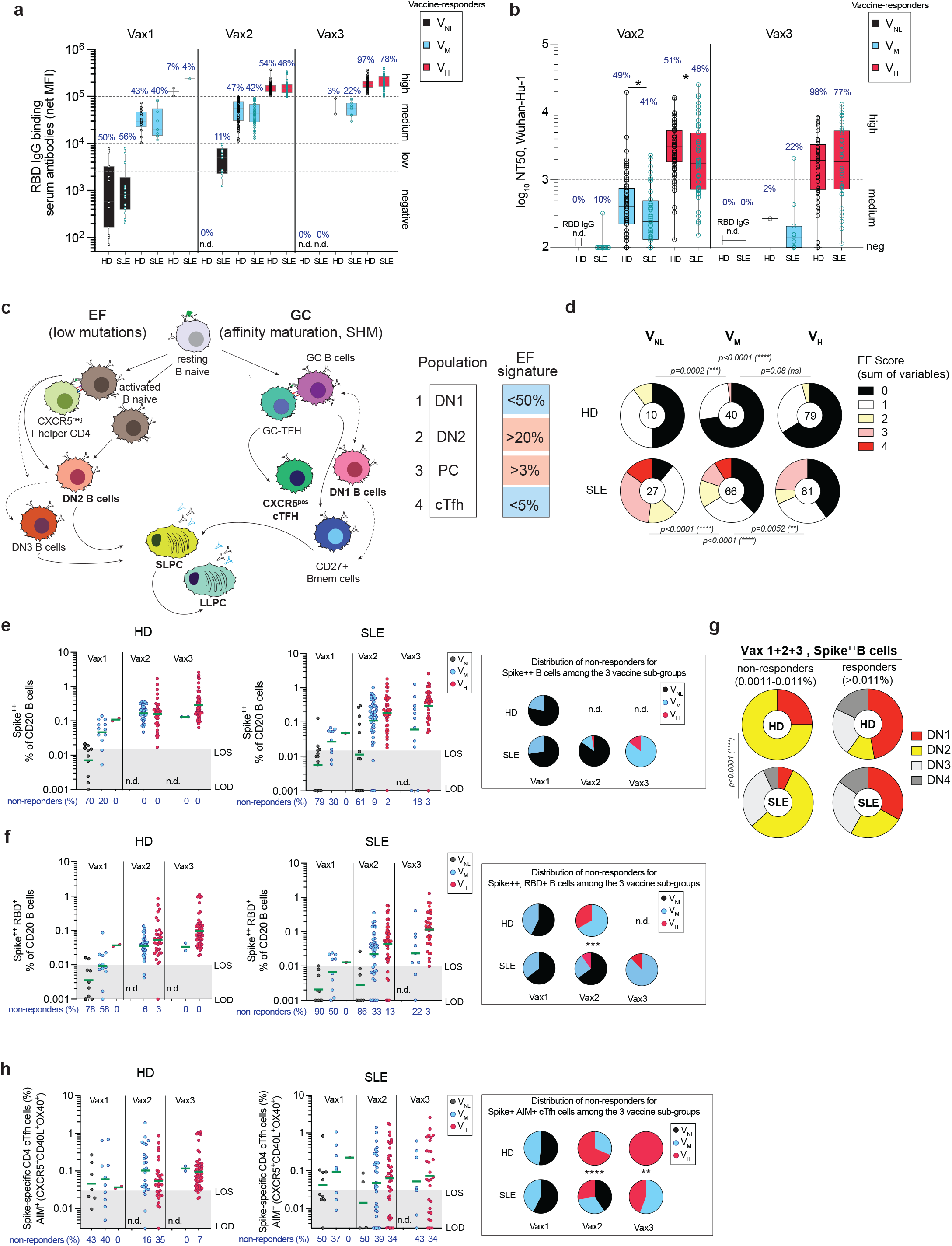
Characterization of our cohort of vaccinees based on vaccine responsiveness and EF-like signature associated. **a**, RBD IgG mean values showed based on the classification of 3 groups: negative/low (V_NL_, 0-10k MFI values), medium (V_M,_ 10k-100k), and high (V_H,_ >100k), and proportion of responders showed for each group and time-point. **b**, Plasma log_10_ NT50 values detected for the pseudoviral neutralization of WA.1 strain and based on the 3 groups of vaccine reactiveness as in (**a**). The proportion of responders showed for each group and time-point and statistical Mann-Whitney U test was applied to compare the groups in HD and SLE. **c**, Simple model showing the proposed B cell differentiation alongside the EF or GC pathways. Variables (cell population frequency) used to mimic a circulating B and T cells EF-derived signature based on reduced frequency of CD21^+^CD11c^-^ DN1 (<50%) and CXCR5^+^ cTfh (<5%) and increased of CD21^-^CD11c^+^ DN2 (>20%) and CD20^neg^CD38^hi^ PC (>3%). **d**, Variables related to the “EF signature” were scored and summed in each sample tested for V_NL_, V_M,_ and V_H_ groups in HD and SLE. Intra-cohort and inter-cohort statistical comparisons were performed with the Chi-square test. **e-h**, Cellular analysis of spike reactive cells based on vaccine-responsiveness and frequency of non-responders at each vaccination dose. Frequency of **e**, spike^++^ B cells, **f**, spike^++^, RBD^+^ B cells, and **g**, DNs reactivity among non-responders (analysis on spike signals lower than baseline, or LOS) and **h**, AIM^+^ cTfh. Each dot represents a sample tested at the indicated time point. The frequency of the samples identified as “non-responders” (based on a LOS threshold of the pre-pandemic and pre-vaccination samples) is shown for each group and represented in the pies for the distribution among each vaccine-responsiveness group. The Chi-square was used for statistical comparisons. n.d.=not detected. When indicated, LOD=limit of detection set to logarithmic 0.001 for B cells and 0.003 for T cells. LOS=limit of sensitivity, based on median values of baseline + 2 x SD.

We asked whether the qualitative assessment of the serological reactivity was correlated to the magnitude of antigen-specific memory B and T cells. A strong correlation was detected between negative and low antigen-specific B cell responses and the group of V_NL_ (Fig. 5e, f). Similarly, a reduced or absent T cell reactivity to the spike was also associated with low antibody responses and significantly defined the low AIM^+^ cTfh reactivity in the SLE receiving recall doses (Fig. 5h, Extended Data Fig. 9 f, g). Notably, the lack of immune memory responses in the B and T cell compartments was uniquely present in a group of V_NL_ SLE at Vax2. As observed for the general EF signature in poor responders, we also detect a more non-conventional memory and EF-driven antigen-specific B cell differentiation, with DN2 reactivity enriched in non-responders’ samples (Fig. 5g).

### BAFF inhibition in SLE is associated with high EF scores and low vaccine responses

We next investigated cellular variables within the different SLE treatment groups. Integrating our immunophenotyping post-vaccination data through principal component analysis (PCA), segregated the HD and SLE cohorts based on underlying immunological-associated features (Fig. 6a and Extended Data Fig. 10a). The results indicate that general immune cell composition remains distinct between SLE and HD cohorts throughout the vaccine-mediated responses. Overlaying patient metadata onto the PCA revealed that, among several classes of drugs administered to our cohort of SLE, subjects receiving anti-BAFF therapy (Belimumab, BLM) were the most distant group from both HD and SLE patients on other class-of-treatment (Fig. 6a, Extended Data Fig. 10b and Table 2). Immune profile comparisons of our vaccinated HD and SLE patients sub-grouped by treatment class confirmed that unique EF signatures were globally associated with the SLE diagnosis. Features included a stronger EF B and T cell signature, expansion of the activated naïve, DN2 and antibody-secreting cell in the B cell compartment and reduced circulating cTfh cells (Extended data Fig. 10 c-h).

**Fig. 6.**
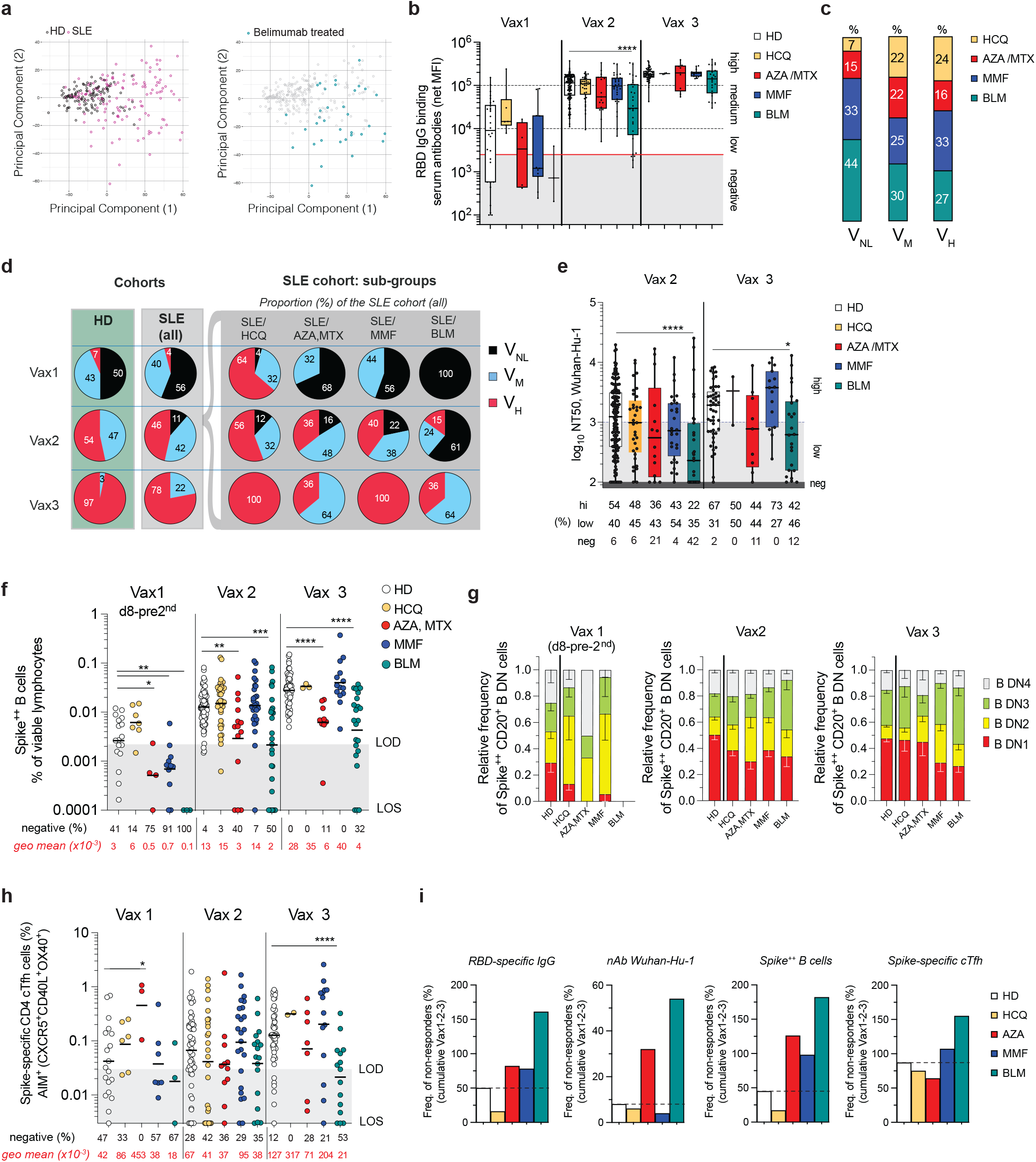
SLE treatment impact on vaccine-mediated responses and enrichment of poor responders in the SLE/BLM group. **a,** Unsupervised PCA analysis showing separation of HD and SLE based on immunophenotypic variables **b**, Luminex-based detection of RBD IgG binding serum antibodies (net MFI values) in the cohort of HD and sub-groups of SLE based on their group of treatment. **c**, Proportion of SLE group based on their treatment among the 3 groups of V_NL_, V_M,_ and V_H_ vaccine-responders. **d**, Table with pies showing the percentage of V_NL_, V_M,_ and V_H_ for HD and SLE, and the relative proportion of each SLE/treatment responsiveness of the total SLE cohort. **e**, Neutralizing titers showed based on the SLE group of treatment for Wuhan-Hu-1 (WA.1) strain and distribution of responders shown as a frequency of the total samples for each column. **f**, Cellular analysis of spike reactivity for spike^++^ B cells calculated as the frequency of viable cells and **g,** the relative frequency of spike reactive DN B cell populations for HD and SLE sub-groups. **h**, AIM^+^ cTfh cells shown for each Vax and SLE subgroups. **i**, Distribution of non-responders calculated as the cumulative percentage (%) of negative values for each indicated category, summarized for Vax 1-3 (RBD-specific IgG, spike B and cTfh) and Vax 2-3 (nAb). Each dot represents an individual sample. Statistical comparison between HD and SLE sub-groups per each vaccine dose performed with unpaired, Mann-Whitney U test. When indicated, LOD=limit of detection set to logarithmic 0.001 for B cells and 0.003 for T cells. LOS=limit of sensitivity, based on median values of baseline + 2 x SD.

Overall, the multivariate analysis suggests that a combination of both disease-associated immune phenotype characteristics, but also lupus-specific medications might distinguish vaccine-mediated immune responses.

Therefore, we sought to explore whether there was a correlation between poor vaccine responses and specific SLE treatments. Humoral and cellular responses were diverse and changed dynamically in each SLE sub-group. We observed that the sub-group of SLE/BLM was enriched in the category of poor vaccine responders (V_NL_), while the other sub-groups (Mycophenolate Mofetil (MMF), Azathioprine (AZA), or Methotrexate (MTX)) were similarly distributed among the 3 vaccine groups, with the Hydroxychloroquine (HCQ) sub-group more represented by good vaccine responders (Fig. 6b-d). Total IgG specific RBD levels did also reflected the nAb potency and breadth of the SLE sub-groups (Fig. 6e and Extended Data Fig. 10i). Additionally, administration of a high dose of glucocorticoids (at doses higher than the equivalent of 10 mg/day of prednisone) have also been associated with diminished antibody response in rheumatic diseases^59^, with doses of *≥*20 mg/day considered as strong immunosuppressive during vaccination^60^. In our study, the SLE patients that received steroids (49% of the SLE cohort) were treated with less than 20 mg/day of prednisone or equivalent drugs (range of 1-15 mg/day, median 5± 3 SD); while their responses upon priming (Vax1) were lower than HD and SLE subjects not receiving any steroids, the inclusion of steroids at Vax 2 and 3 did not modify the response relative to groups without steroids (data not shown).

Within poor responders, a general reduction of total circulating B cells (Extended Data Fig 10c) correlates with reduced vaccine responses. Cellular mediated responses were largely unaffected in the HCQ patients, but all SLE were characterized by the enrichment of non-DN1 spike-reactive B cells, partially and selectively rescued by subsequent doses (Fig. 6g). Of the B cell spike reactivity, a general reduction of conventional CD27^+^ memory B cells (accompanied by increased DN B cells) was observed in the SLE, but distinguished the BLM group whom had a greater representation of this CD27^+^ compartment, which is largely unaffected by this treatment^39^ (Extended Data Fig. 10j). T cells responses were broadly reduced in SLE patients, with a more dramatic and general loss of CD8 reactivity against the spike (Extended Data Fig. 10k), and low CD4s and memory cTfh responses in the poor B cell responder groups (Extended Data Fig. 10k and Fig. 6h).

Overall, the most dramatic reduction and lack of vaccine efficiency, measured as a combined activity of serological and cellular mediated responses, was observed in the SLE subjects that were under anti-BAFF treatment at the time of initial vaccination, presenting with the highest enrichment of non-responders (Fig. 6i). Notably, and in keeping with its preferential impact on naïve B cell activation, all BLM-treated patients failed to respond to the priming dose (Fig. 6f) and were characterized by a significant reduction of potency and breadth of neutralization (Fig. 6e, Extended Data Fig. 10i), associated with reduced magnitude of memory antigen-specific B cells (Fig. 6f) and spike-specific cTfh responses (Fig. 6h).

## Discussion

The efficacy of SARS-CoV-2 mRNA vaccines to protect against COVID-19 infection, in particular severe infections and deaths, is well documented in the general population. Protection is mediated by both antibody production, mostly neutralizing antibodies, and T cell responses. Optimal vaccine responses also induce the generation of long-lived plasma cells capable of sustaining antibody responses for years and memory B cells. While pre-existing antibodies serve to prevent or minimize infections, memory B cells ensure quick responses to a new infection that can adapt to new variants. All these immunological functions can be readily measured through serological testing and cellular assays to ascertain the frequency of antigen-specific B cells and T cells elicited by vaccination. While correlates of protection are still under investigation ^17^, anti-RBD IgG and neutralizing antibodies (nAb) have been proposed as predictors of vaccine efficacy and protection from viral infections. Cellular memory responses have also been proposed as the coordinated activity of both antigen-reactive memory B and T cells may correlate with high levels of anti-RBD IgG, virus neutralization, and cell-mediated virus clearance^61^.

The clinical efficacy and immune correlates have been amply tested in the healthy population as well as in cancer patients^44,62^ and patients with solid organ transplantation^63-68^. However, the humoral and cellular responses to the mRNA vaccines are poorly understood in patients with SLE in whom both components can be compromised by the autoimmune disease itself as well as multiple immunosuppressive treatments. Indeed, extant studies in SLE have been largely limited to antibody testing and performed in relatively low numbers of patients with under-representation of severe Black SLE^24,69^.

Our study represents the first *in-depth* antibody and cellular analysis of a large cohort of SLE with a majority of Black patients (n=79; 68% Black), that received only a primary vaccine series (Vax1+2) or an additional third-dose booster (Vax3) of the SARS-CoV-2 mRNA vaccines coding for the WA.1 spike. Globally, SLE subjects showed reduced seroconversion rates and lower mean titers of anti-RBD IgG for up to 6 months after the second dose, while HD seroconverted at a higher rate after priming and fully after Vax2. Of note, in the HD, the amount of circulating anti-RBD IgG correlated strongly with inhibition of RBD binding to ACE2, a surrogate of potent neutralizing antibodies capable of blocking viral entry^70^. In contrast, the SLE group displayed a generalized reduction of ACE2 blocking activity that was uncoupled from anti-RBD IgG titers. In harmony with the lower competitiveness, the neutralizing power and breadth of neutralization of the SLE plasma samples were significantly lower than those of the control group, even after full seroconversion. While the mechanistic basis of these qualitative antibody defects remains to be determined, we propose that these features could be explained by defective affinity maturation due to aberrant GC activity and/or dominant extra-follicular activity^71-73^. Our findings warn against assessing the mRNA vaccine efficacy on some vulnerable populations simply based on the total anti-spike or anti-RBD IgG titers. Also of interest, is the slower decline in neutralizing antibody titers observed post-booster in SLE patients, a finding that could be explained by higher production of long-lived plasma cells in this disease^74^.

Most vaccine studies are limited to antibody measurements although a small number have also analyzed B cell memory responses upon influenza vaccination and more recently, upon mRNA SARS-CoV-2 boosters^12,50,51,75-77^. Recent studies have used lymph node fine needle aspirates (FNA), to demonstrate prolonged persistence of germinal center reactions and the competition between pre-formed memory B cells and new naïve arrivals in influenza recall responses and upon SARS-CoV-2 mRNA vaccination and monovalent booster^6,13,78,79^. FNA has also been used to demonstrate defective GC reactions and memory B cell formation in kidney transplant recipients^63^. By and large, antigen-specific memory B cells were identified within the conventional CD27^+^ population or adjudicated without discriminating phenotypic markers^75^. One study used additional markers to show that CD11c^+^ activated cells increased after each vaccine dose and declined significantly faster than CD11c^-^ CD27^+^ memory B cells. However, long-term maintenance of memory after boosting was not assessed^12^.

As new B cell populations continue to be identified, SARS-CoV-2 vaccination offers a unique opportunity to understand human primary responses and how memory is initiated upon the initial priming and then diversified with successive boosters. We had previously showed that in severe Black SLE, and despite strong pre-existing autoimmune memory, disease flares are characterized by an influx of newly activated naïve cells whose differentiation through an extrafollicular pathway generate a large fraction of the antibody secreting cells (ASC), expanded during active disease^35,48^. Those studies identified effector DN2 B cells downstream of activated naïve B cells. Termed DN2 cells owing to the absence of IgD and CD27 and the expression of CD11c and T-bet, these effector cells are epigenetically poised in SLE to differentiate into ASC^80^. The application of this information allowed us to demonstrate that this pathway is highly activated within the first few days of severe COVID-19 infections leading to the generation of both anti-viral and autoreactive antibodies^36,49,81^ and enabled us to interrogate the response to SARS-CoV-2 vaccination in SLE.

To the best of our knowledge, the present study provides the first comprehensive characterization of effector and memory responses to COVID-19 vaccines and the first of this nature in patients with SLE. Enabled by the analysis of antigen-specific B cells during the effector and memory phases of primary and recall responses through three vaccine administrations over 18 months, one notable result is the demonstration of the contribution of DN2 cells to both, the effector and memory responses. This finding indicates the limitations of ascribing memory function on the basis of phenotypic markers that are shared by effector cells, including loss of CD21 and expression of CD11c. Moreover, it strongly suggests the ability of human EF responses to generate a separate memory cell compartment, a phenomenon already established in mice^81^. However, final proof of this concept will necessitate additional longitudinal studies with BCR repertoire linkage. In keeping with the accentuation of the EF pathway in severe SLE and its potential contribution to a memory compartment, DN2 cells dominated the effector phase of the priming response and remained a large component of spike-specific B cells several months after boosting in the absence of intervening infection. Consistent with a similar degree of heterogeneity, an equivalent pattern was followed by DN3 cells, another component of the EF response that we and others have found to be expanded in SLE and severe COVID-19 infection^36,53,82^. In contrast, DN1 cells presumed to represent early GC memory^35^, dominated normal responses across all vaccination timepoints. Of great interest, within SLE patients, a high EF index derived from the combination of DN1, DN2, PC and cTfh values, strongly correlated with lower antibody responses. Combined, our data are consistent with a model in which an EF B cell SLE endotype would promote decreased antibody responses with reduced neutralization activity presumably on the basis of defective affinity maturation. They also support the functional consequence of B cell endotypes and their heterogeneity in SLE^34,53^.

Also indicative of the SLE milieu with high levels of type II interferon in association with enhanced EF responses^83^, antigen-reactive B cells in SLE patients were significantly enriched in CXCR3-bearing cells^83-85^. Notably, the priming response of HD to the first vaccine dose was significantly enriched, relative to SLE, in CCR6^+^ cells devoid of CXCR3, with these cells becoming the majority of HD memory cells persisting for months after boosting. The HD pattern could promote an enhanced GC origin of memory cells in normal responses and a heightened predisposition of pre-formed memory cells to participate in subsequent GC reactions^86-88^. In contrast, the SLE profile could promote relocation of tissue CXCR3^+^ memory cells to sites of viral re-infection^89^. An interesting observation on the B cell compartment showed an opposite trend of durability for the secreted antibodies and the magnitude of spike-reactive cells in the SLE cohort, suggesting that in the latter, prolonged plasma cell responses and circulating Ab can interfere with memory B cell formation.

In addition to disease-related determinants, the response to vaccination in SLE is also modulated by treatment. Previous studies, restricted to antibody assessment, have predictably shown that B cell and plasma cell targeting agents can dampen antibody-mediated responses and the global B cell pool^44,56,62^. Other immunomodulatory drugs like MMF, have been shown to reduce vaccine efficacy in some vulnerable populations such as autoimmune rheumatic^22,24,69^ or solid organ transplant recipients^63-68^. In our SLE cohort, we did also observe reduced seroconversion associated with MMF treatment. Yet, in contrast with kidney transplants, the second and third doses were able to rescue the initially deficient responses^67^. This discrepancy can be due to both disease-specific features, the dose of MMF administered, and co-treatments with other potent immunosuppressing drugs.

Notably, in SLE the most profound and persistent deficit across the immune landscape was observed in patients treated with Belimumab (BLM), including the poorest total antibody and neutralization titers. Indeed, as predicted by the preferential role of BAFF in the survival of naïve B cells and the known global naïve B cell lymphopenia induced by its blockade^90-92^, BLM treatment correlated with highly defective antigen-specific B cell primary responses and, presumably as a consequence of defective priming, with the smallest B cell memory compartment even after boosting. Within that reduced memory pool, BLM-treated patients displayed the highest frequency of CD27^+^ memory cells, a feature consistent with the established BAFF-independent generation and survival of GC-dependent memory cells^93,94^. Finally, BLM-treated patients also had the lowest levels of spike-reactive cTfh cells. Combined, our data suggest that the role of BAFF in the generation of cTfh cells and in the affinity maturation of GC B cells, could explain how BLM treatment results in poor antibody ACE2 competitiveness, neutralization and breadth potency^95,96^.

In sum, we demonstrate quantitative and qualitative B cell, T cell and antibody defects in the response of SLE patients to mRNA vaccination. The investigation of these patients sheds light into the B cell biology underlying the autoimmune process and contributes original insight into the functional consequences of disease-related abnormalities in vaccine responses. Similarly, the study provides new information regarding the impact of different treatments on B cell responses. Noteworthy, it demonstrates the role of BAFF, a cytokine of central significance in SLE, on different components of the B cell, T cell and antibody response.

Globally, the knowledge acquired in this study, should form the basis for new strategies to enhance the efficacy and durability of mRNA vaccinations, including modifications of vaccine dosing and timing and modulation of the magnitude and/or timing of the immunosuppressive agents. Specifically, we would suggest that the initiation of BLM should be delayed until the first two doses have been administered in order to optimize priming and development of the early memory pool. The same benefit could be promoted by a two-months “vacation” in patients already receiving BLM. More frequent dosing over the first three months of the vaccination schedule could also overcome the overall deficit in seroconversion observed in SLE while maximizing the ability of these patients to generate sustained antibody responses through the generation of long-lived plasma cells. Whether newer therapies such as interferon blockade will impact the priming and development of memory responses^97,98^ and the balance of long-lived and short-lived plasma cells^99,100^ will require additional studies of the increasing number of patients treated with this recently approved modality.

## Supporting information

Ext Data

## Data Availability

Data available upon request.

## Methods

### Human participants

All research was approved by the Emory University Institutional Review Board (Emory IRB nos. IRB000057983 and IRB000058515) and was performed in accordance with all relevant guidelines and regulations. Blood draws were obtained after the subjects’ written informed consent. Healthy individuals (n = 64) and patients diagnosed with lupus (SLE) (n = 79) were recruited from Emory University Hospital, Emory University Hospital Midtown, and Emory Grady (all Atlanta, USA). All HD and SLE demographics and SLE medications are listed in Table 1 and Table 2. Study data were collected and managed using REDCap electronic data capture tools hosted at Emory University. All subjects enrolled in this study were considered naïve-to-SARS-CoV-2 infections based on the absence of self-reported symptoms and/or negative PCR tests. All tested samples were screened for the absence of SARS-CoV-2 Nucleocapsid (N) reactive IgG and N-positive samples were excluded from the study (Extended Data Fig. 2a, b).

### Peripheral blood mononuclear cell (PBMC) isolation and sera and plasma collection

Peripheral blood samples were collected in Green-top (Vacutainer) sodium heparin tubes. Red-top tubes (Vacutainer) were centrifuged for 10 min at 800 g to collect sera. Undiluted plasma was collected after centrifugation of blood for 10 min at 500 g. Sera and plasma aliquots were stored at -20 degrees Celsius. Next, the blood was diluted (1:2) with PBS and PBMCs were isolated by Ficoll density gradient centrifugation at 1000g for 10 min. PBMCs were washed twice with R10 complete medium and ACK lysed. Viability was assessed using trypan blue exclusion, and live cells were counted using an automated hemocytometer. Frozen cells were stored at -80 degrees Celsius in FBS 10% DMSO.

### Flow cytometry

Frozen vials of PBMCs were thawed, washed, and centrifuged at 800 g for 5 min, and then resuspended in warm R10 complete medium at a final concentration of 20 x 10^6^/ml. Cell suspensions were divided into 3 parts to perform 1) AIM assay (∼1 x 10^6^), 2), B cell tetramer staining (∼4-8 x 10^6^), and 3) T cell staining (∼2-4 x 10^6^). The mix of the diluted antibodies was prepared in staining buffer (PBS 2% FCS, 1 mM EDTA) supplemented with Super Bright Complete Staining Buffer and added to the samples in a 96-well round bottom plate, at a final volume of 50 μl/well. After each step, the abs staining was blocked with a wash in staining buffer (∼100 μl) and centrifugation of the plate at 800 g, for 5 min. For the B cell immunophenotype, after incubation with the tetramers for 1 hour at 4 degrees Celsius, cells were washed and stained with a mix of anti-Igs (IgD + IgM + IgA + IgG) for ∼15 min at RT, in the dark. The cells were then incubated with the cocktails of the remaining abs for ∼40 min at RT, in the dark. After washing and centrifugation, cells were stained with L/D Zombie NIR (diluted at 1:500 in PBS) for 10 min at RT, in the dark. Similarly, for the immunophenotype of T cells, samples were stained with the abs mix for ∼40 min at RT, in the dark, and viable cells were detected with L/D Zombie NIR. Cells were resuspended in ∼180 μl of staining buffer, kept on ice, and immediately analyzed on a 5L Cytek Aurora flow cytometer using Cytek SpectroFlo software.

### Detection of tetramer-binding B cells

Antigen-specific B cells were detected using tetramer-probes. Biotinylated spike and RBD proteins (R&D Systems) were multimerized with fluorescently labeled streptavidins (SAs) for 1 hour at 4 degrees Celsius. Full-length spike protein was mixed with SA-BV421, using ∼100 ng of spike and ∼20 ng of SA for each sample (∼4:1 molar ratio). RBD was mixed with SA-AF647, using ∼25 ng of RBD and ∼12.5 ng of SA (∼4:1 molar ratio). The SA-PerCP was used as a decoy probe. After 1 hour, all tetramers were mixed in free D-biotin (5 μM, Avidity LLC) to minimize cross-reactivity. PBMCs were incubated for 1 hour at 4 degrees Celsius with the mix of tetramers, washed and stained with surface markers, and labeled for viability with L/D Zombie NIR (1:500) for ∼10 min prior to being acquired with the 5L Aurora (Cytek).

### AIM assay

Activation Induced Markers (AIM) assay was performed as follows. Briefly, PBMCs were counted and adjusted to 20 x 10^6^ cells per ml in complete medium (RPMI-1640 5% Human Sera (AB) supplemented with Glutamax, N-EAA, NaPy, and Pen-Strep). Antigen-specific T cells responses were assessed by incubating 1 x 10^6^ cells with MPs “mega-pools” of peptides (15-mers overlapping by 11aa) encompassing the sequenced of human pathogens, SARS-CoV-2 Wuhan-Hu-1 full spike (aa 5-1273, Miltenil) or HA (H1N1, strain swl A/California/04/2009, Miltenyl). Cells were incubated in a 96-well U-bottom plate in a final volume of 200 μl. To detect extracellular CD40L expression, cells were pre-incubated with an anti-human CD40 antibody (Miltenyl Biotech, pure-functional grade, Catalog #130-094-133) used at 0.5 μg/ml, for 15 minutes at 37 degrees Celsius prior to stimulation. Cells were then stimulated with each MPs of spike or HA (used at 1 μg/ml). Polyclonal stimulation of cells was assessed with TCR triggering via SEB (used at 1 μg/ml) and used as a positive control. Unstimulated cells at the same density were incubated in complete media and used as background signals for the quantification of AIM^+^ cells. Cells were stimulated for ∼24h before washing and staining for flow cytometry. Cells were acquired on a 5L Aurora (Cytek). The data in the graphs are shown as background subtracted (controls) values and negative values are shown as 0.003 in the log_10_ scale graphs.

### Carbodiimide coupling of microspheres to SARS-CoV-2 antigens and Luminex proteomic assays for measurement of anti-antigen antibody

This analysis was carried out as previously described^40^. Briefly, five SARS-CoV-2 proteins were coupled to MagPlex Microspheres of different regions (Luminex). The N protein (catalog no. Z03480; expressed in *Escherichia coli*) and the S1-RBD (catalog no. Z03483; expressed in HEK293 cells) were purchased from GenScript. The S1 domain (aa 16--685; catalog no. S1N-C52H2; expressed in HEK293 cells), the S1-NTD (aa 13--303; catalog no. S1D-C52H6; expressed in HEK293 cells), and the S2 domain (catalog no. S2N-C52H2; expressed in HEK293 cells) were purchased from AcroBiosystems. Each protein was expressed with an N-terminal His6-tag to facilitate purification, >85% pure and appeared as a predominant single band on SDS-PAGE analysis. Coupling was carried out at 22 °C following standard carbodiimide coupling procedures. Concentrations of coupled microspheres were confirmed by Bio-Rad T20 Cell Counter. Approximately 50 μl of coupled microsphere mix was added to each well of 96-well clear-bottom black polystyrene microplates (Greiner Bio-One) at a concentration of 1,000 microspheres per region per well. All wash steps and dilutions were accomplished using 1% BSA, 1× PBS assay buffer. Sera were assayed at 1:500 dilutions and surveyed for antibodies against N, S1, S2, NTD or RBD. After a 1-h incubation in the dark on a plate shaker at 800 r.p.m., wells were washed five times in 100 μl of assay buffer, using a BioTek 405 TS plate washer, then applied with 3 μg ml−1 PE-conjugated goat anti-human IgA, IgG and/or IgM (Southern Biotech). After 30 min of incubation at 800 r.p.m. in the dark, wells were washed three times in 100 μl of assay buffer, resuspended in 100 μl of assay buffer and analyzed using a Luminex FLEXMAP 3D instrument (Luminex) running xPONENT 4.3 software on the Enhanced PMT setting. MFI using combined or individual detection antibodies (anti-IgA, anti-IgG or anti-IgM) was measured using the Luminex xPONENT software. The background value of assay buffer was subtracted from each serum sample result to obtain MFI minus background (MFI-B; net MFI).

### RBD competitive ELISA assay

The ELISA plates 96-well half-area (Corning, #3690) were pre-coated overnight with 2 μg/ml of recombinant human-ACE2 in PBS, at 4 degrees Celsius. Plates were blocked with Casein 1% for 30 min at 37 degrees Celsius. Serially diluted plasma/sera were mixed with RBD mouse Fc-tagged antigen (Sino Biological, final concentration 20 ng/mL) for 30 min at 37 degrees Celsius, and the mix was then added to the plates, for 30 min at 37 degrees Celsius. The plates were washed with PBS 0.1% Tween^20^ and RBD binding was revealed using secondary goat anti-mouse IgG (Southern Biotech #1031-05, 1:5000 in PBS). Plates were incubated for 30 min at RT and, after washing, 40 μl of TMB substrate was added and developed plates were blocked with 40 μl of Stop Solution and read at 405 nm with an ELISA reader. The percentage of inhibition was calculated as follows: (1-(OD sample - OD neg ctr)/(OD pos ctr - OD neg ctr)]) x 100.

### Pseudoviral *in vitro* neutralization assay

Neutralization activities against SARS-CoV-2 WT (Wuhan Hu-1), Delta (B.1.617.2) and Omicron BA.1 strains were measured in a single-round-of-infection assay with pseudoviruses, as previously described^44^. Briefly, to produce SARS-CoV-2 WT, Delta and Omicron pseudoviruses, an expression plasmid bearing codon optimized SARS-CoV-2 full-length S plasmid (parental sequence Wuhan-1, Genbank #: MN908947.3) was co-transfected into HEK293T cells (ATCC#CRL-11268) with plasmids encoding non-surface proteins for lentivirus production and a lentiviral backbone plasmid expressing a Luciferase-IRES-ZsGreen reporter, HIV-1 Tat and Rev packing plasmids (BEI Resources NR-53818) and pseudoviruses harvested after 48 hours of post transfection and performed titration. Pseudoviruses were mixed with serial dilutions (1:50 to 1:328,050) of plasma, incubated 1 hour for the reaction at 37 degrees Celsius, 5% CO2 incubator, and then added to monolayers of ACE-2-overexpressing 293T cells (BEI Resources NR-52511), in duplicate. Forty-height hours after infection, cells were lysed, luciferase was activated with the Luciferase Assay System (Bright-Glo Promega), and relative light units (RLU) were measured on a synergy BioTek reader. Statistical analysis was performed using the software GraphPad prism 9.0 for determination of ID50 values through a dose-response curve fit with non-linear regression.

### Principal Component Analysis

PCA plots were generated in R (v3.6.2; release 12 Dec 2019) using the ‘ggbiplot’ library. Custom plotting was performed using the ‘ggplot2’ library for base analysis, and then post-processed in Adobe Illustrator.

### Half-life/Durability analysis

Immune responses, starting 42 days post 2^nd^/3^rd^ mRNA shot, were analyzed using linear mixed effects models with log immune response as the dependent variable and time as the independent variable. Models for SLE patients, healthy donors and combined included population-level fixed effects and individual-level random intercepts. Statistical significance of fixed effect coefficients was assessed via Wald test. Those with maximum response ≤ the 95^th^ percentile of baseline and pre-pandemic samples and data associated with unexpected > 2-fold increases in NTD IgG were excluded. Models were fit using lmer function from lme4 package and R programming language.

### Statistical analysis

Statistical analysis was carried out using Prism v.9.5.1 (GraphPad Software). For each experiment, the type of statistical testing, summary statistics and levels of significance can be found in the figures and corresponding legends. *\*P < 0.05, **P < 0.01, ***P < 0.001, ****P < 0.0001*.

## Acknowledgments

We thank all the donors for their generosity in participating in and making this study possible. We thank the nurses, staff, and providers in the Emory Clinics and Hospitals in Atlanta (GA).

We thank the members of the Sanz and Lee labs for helpful discussions. We would like to also thank the Sanz/Lee clinical coordination and sample processing teams for sample identification, scheduling, and collection. We thank Dr. Kevin S. Bonham for helpful discussions on biostatistical analyses. Finally, we would like to thank Drs. Davide Corti, Luca Piccoli and Jessica Bassi for helping with the ELISA blocking protocol delineation.

This work was supported by National Institutes of Health grants: U54-762 CA260563-01 Emory SeroNet (I.S., F.E.L.), U19-AI110483 Emory Autoimmunity Center of Excellence (I.S.).

## Author contributions

C.E.F and I.S. conceived and directed the study. C.E.F. processed samples, performed most experiments and data analyses. M.C.W. performed PCA analyses. H.A. performed half-life and durability of antibody and cellular-mediated fold-change responses comparisons between cohorts. T.T.P. helped in performing flow and ELISA assays, and samples’ organization. M.C.R. and F.E.L. provided data from healthy controls tested for viral antigens. N.C. and S.C. performed pseudoviral *in vitro* tests with plasma samples. F.A.A., N.S.H., A.M.P, and H.Q. performed serum screening against viral antigens. S.Y.U. and A.K. conducted chart review, identified patient samples for study inclusion and provided donors’ information. S.K. and F.E.L. coordinated healthy donor samples’ processing and the biobank repository. W.C., A.S.N, J.D.R., R.A. participated in the manuscript review. C.E.F. and I.S. wrote the manuscript, with all authors providing editorial support.

## Extended Data Figure legends

**Extended Data Fig. 1 | Study cohorts’ samples and timeline of samples’ collection.**

**a**, Study design, vaccine administration scheme, and time points collected after SARS-CoV-2 mRNA vaccination for healthy controls and patients with SLE. **b**, Diagrams showing the longitudinal collection of blood draws from HD and SLE enrolled in the study (circles) and pre-pandemic controls (Pre-CoV, squares). The SLE cohort is grouped for the main treatment by colored circles: Untreated (none, black), Hydroxychloroquine (HCQ, yellow), Azathioprine (AZA, red), Methotrexate (MTX, dark red), Mycophenolate Mofetil (MMF, dark blue), Belimumab (BLM, turquoise).

**Extended Data Fig. 2 | High-dimensional flow cytometry characterization of B and T cells. a,** Representative gating strategy for the quantification of CD19^+^ B cells, their subsets, and tetramer-based reactivity detection for spike and RBD by flow cytometry. **b**, Representative gating strategy for quantification of CD3^+^ T cells, and their subsets by flow cytometry.

**Extended Data Fig. 3 | Detection of circulating antigen-specific Igs upon vaccination.**

**a,** Cartoon showing the Luminex bead-based assay. **b,** Nucleocapsid IgG threshold used to define SARS-CoV-2 negative samples. IgM reactivity to the SARS-CoV-2 proteins NTD, S2, S1 and RBD and percentage of responders in **c**, HD and **d**, SLE. IgA reactivity to the SARS-CoV-2 proteins NTD, S2, S1 and RBD and percentage of responders in **e**, HD, and **f**, SLE. Each dot represents a sample tested. Negative values are based on MFI values of time 0 (T0) from pre-pandemic and vaccine baseline samples.

**Extended Data Fig. 4 | Detection of circulating non-RBD targeting Igs upon vaccination.**

Kinetics analysis for (**a**) S1, (**b**) RBD, (**c**) S2, and (**d**) NTD binding IgG. Overlay of geometric mean and pie charts showing the proportion of negative, medium, or high values.

Statistics of the geomean between HD and SLE were performed with U Mann Whitney for comparison of each indicated time. Pie chart comparison statistics performed with Chi-square with Fisher test analysis.

**Extended Data Fig. 5 | Detection of circulating neutralizing Igs upon vaccination.**

**a,** Cartoon showing the steps of pseudoviral *in vitro* neutralization assay. **b,** NT50 values detected from HD and SLE plasma and comparison of all viral strains for each sample. Fold changes show the difference between Delta and Omicron compared to WT values.

**Extended Data Fig. 6 | B cell profile and spike reactivity.**

**a,** Circulating CD19^+^ B cells frequency comparison in our cohorts. **b,** Correlation of CD19^+^ frequency and spike reactive B cells in HD and SLE for each vaccine time point. Blue lines show the range observed in the HD. Pearson analysis was performed for each comparison. Quantification of total (**c**) spike-specific and (**d**) RBD-specific B cells shown as the frequency of viable lymphocytes in the HD and SLE cohorts. Each dot represents an individual sample tested at baseline (d0) and after receiving one (Vax 1), two (Vax 2), or three (Vax 3) vaccine doses. Pie charts comparisons and Chi-square with Fisher’s test are shown. When indicated, LOD=limit of detection set to logarithmic 0.001 for B cells and 0.003 for T cells. LOS=limit of sensitivity, based on median values of baseline + 2 x SD.

**Extended Data Fig. 7 | Ig surface expression of spike^++^ B cells revealed reduced IgG class-switching in SLE.**

**a,** Representative flow detection of Ig surface expression on B cells. **b**, Distribution of IgM, IgG and IgA and Chi-square (Fisher) comparison. Frequency of membrane-bound **c**, IgM, **d**, IgG and **e**, IgA in HD and SLE spike^+^ B cells showed for each vaccine time point as shown in (**b**). Each dot represents an individual sample. The overlay of the median signals for HD and SLE is shown for each Ig tested. One-way ANOVA test is shown within the HD or SLE groups.

**Extended Data Fig. 8 | Analysis of antigen-specific T cell responses.**

**a**, Cartoon showing the AIM assay. **b**, Representative flow plots and gating strategies used to define AIM^+^ T cells and their differentiation. Polarization of AIM^+^ helper T cells in (**c**) total CD4^+^ cells and (**d)** CXCR5 expressing cTfh. **e**, Frequency of AIM^+^ cTfh17 upon Vax1, 2 or 3. **f**, Reactivity towards the HA MP for all tested samples in the AIM assay, and comparison of HD and SLE. **g**, Differentiation of AIM^+^ HA T cells based on CD45RA and CCR7 expression.

**Extended Data Fig. 9 | Clinical, demographics, and immunological features characterizing our two cohorts of vaccinated HD and SLE.**

**a,** Age and race showing the distribution in HD and SLE based on vaccine responsiveness (negative/low V_NL_, medium V_M_, or high V_H_). **b,** Distribution of females (F) and males (M) in our cohorts, based on vaccine responsiveness (negative/low V_NL_, medium V_M_, or high V_H_). **c,** Number of subjects within each group based on age and vaccine responses clustered by age (18-50, >50 years old, yo). **d**, Vaccine responsiveness based on sex (M/F) in our two cohorts. **e**, Correlation of cTfh frequency and B cells subsets frequency (DN1, DN2, ASC, and PC) detected in HD and SLE groups. Pearson analysis was performed for each comparison. **f**, AIM^+^ CD8 and **g**, AIM^+^ CD8 frequencies and percentage of non-responders identified for each vaccine group of V_NL_, V_M,_ and V_H_. When indicated, LOD=limit of detection set to logarithmic 0.001 for B cells and 0.003 for T cells. LOS=limit of sensitivity, based on median values of baseline + 2 x SD.

**Extended Data Fig. 10 | Immunophenotype of vaccinated SLE categorized by treatment.**

**a,** Variance plot driving the PCA analyses. **b,** Simple cartoon showing lupus-associated drugs targeting B cells **and** table showing the main group of SLE divided by treatment as analyzed in this study. **c-h**, Representative high-dimensional flow cytometry characterization of B cell in HD and SLE groups showing the comparison of **c**, CD19^+^ **d**, Naïve B cells, activated Naïve (aN) B cell, Transitional B cells (Btr) and CD21^lo^ Btr. **e**, CD27^+^ conventional memory. **f**, DN B cells and subsets; ratio of DN2:DN1 expressed as log2 value. **g**, IgD^-^ ASC and CD20^-^CD38^hi^ PC. **h**, cTfh and cTfh17. SLE sub-group frequency of each population is compared to the HD group. **i,** NT50 values for Delta and Omicron neutralization observed in HD and sub-groups of SLE. **j**, Relative frequency of spike reactive B cell populations for HD and SLE sub-groups based on CD27 and IgD expression. **k**, Geomean values for spike reactive CD8 and CD4 within the SLE sub-groups. Each dot represents an individual sample. Statistical comparison between HD and SLE sub-groups per each vaccine dose performed with unpaired, Mann-Whitney U test. When indicated, LOD=limit of detection set to logarithmic 0.001 for B cells and 0.003 for T cells. LOS=limit of sensitivity, based on median values of baseline + 2 x SD.

