## Supplementary material for "Poor immunogenicity upon SARS-CoV-2 mRNA vaccinations in autoimmune SLE patients is associated with pronounced EF-mediated responses and anti-BAFF/Belimumab treatment": Ext Data

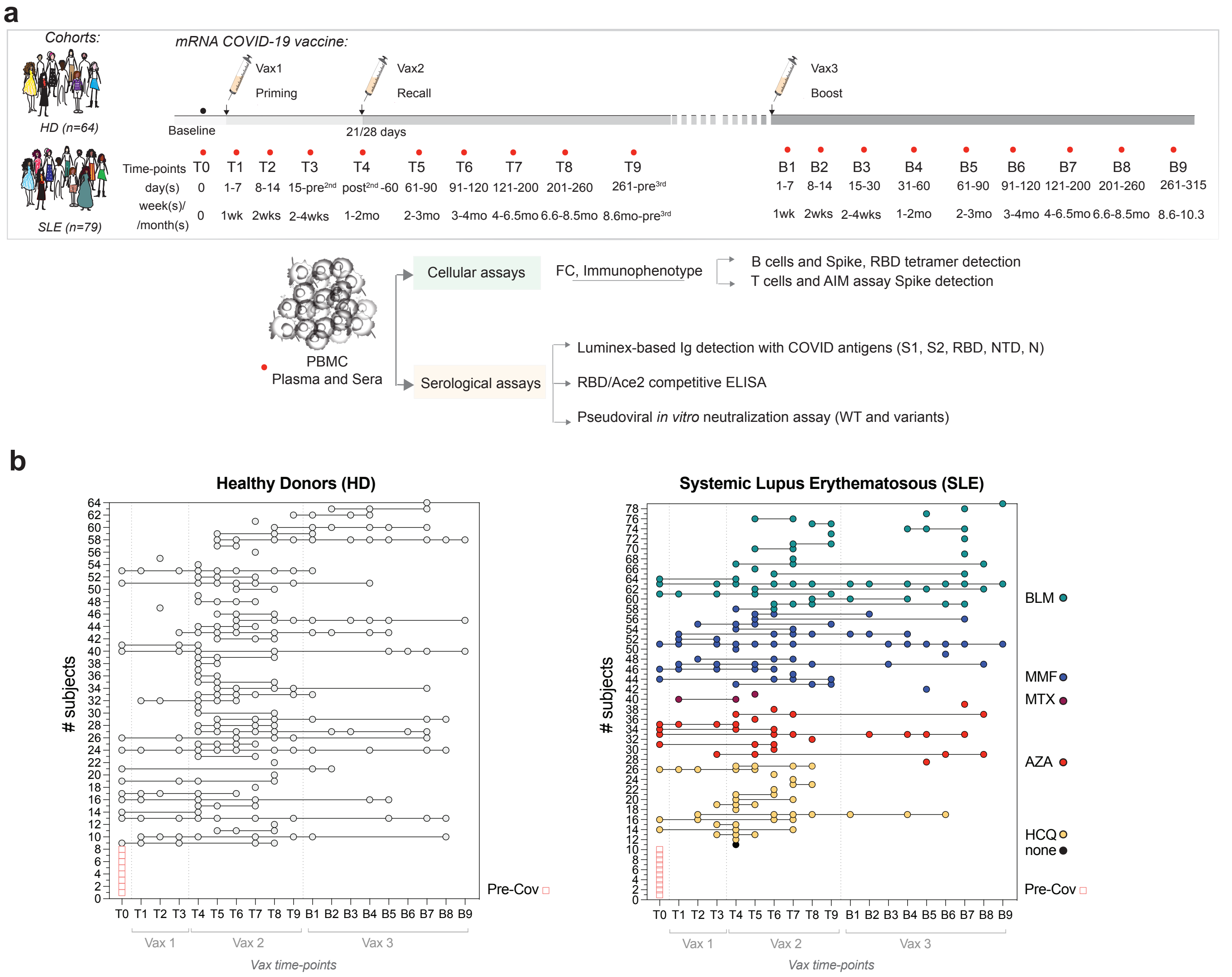

**Extended Data Fig.1 | mRNA SARS-CoV-2 vaccination in HD and SLE.**
**Study design, time-points of samples collection and longitudinal sampling of donors enrolled.**

**a**

High-dimentional Flow cytometry - B cells immunophenotype

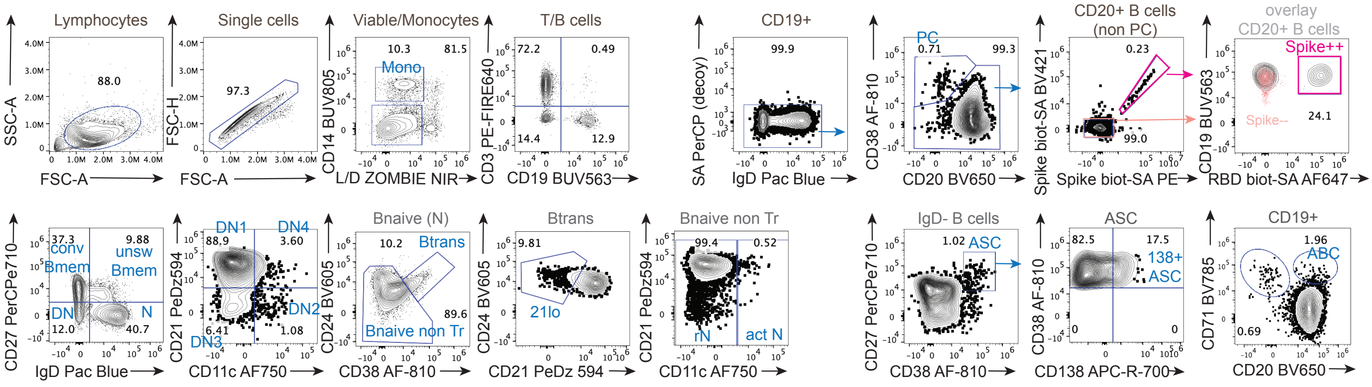

**b**

High-dimentional Flow cytometry - T cells immunophenotype

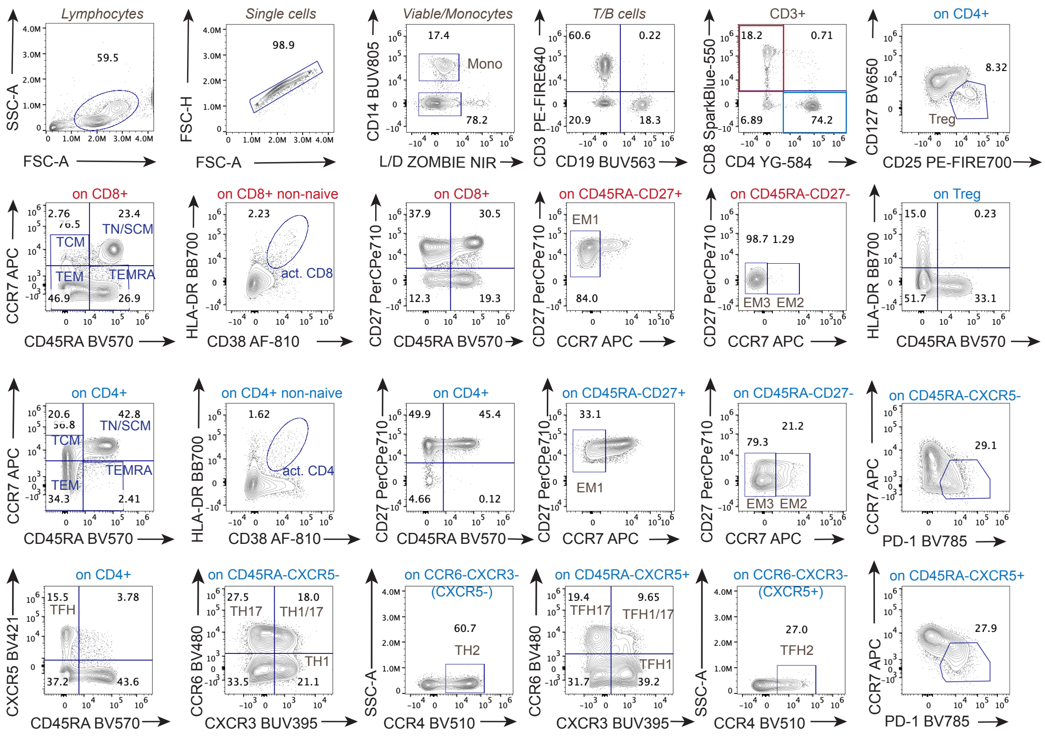

**Extended Data Fig.2 | High-dimentional flow immunophenotyping of B and T cells in the subjects enrolled in the mRNA vaccine study.**

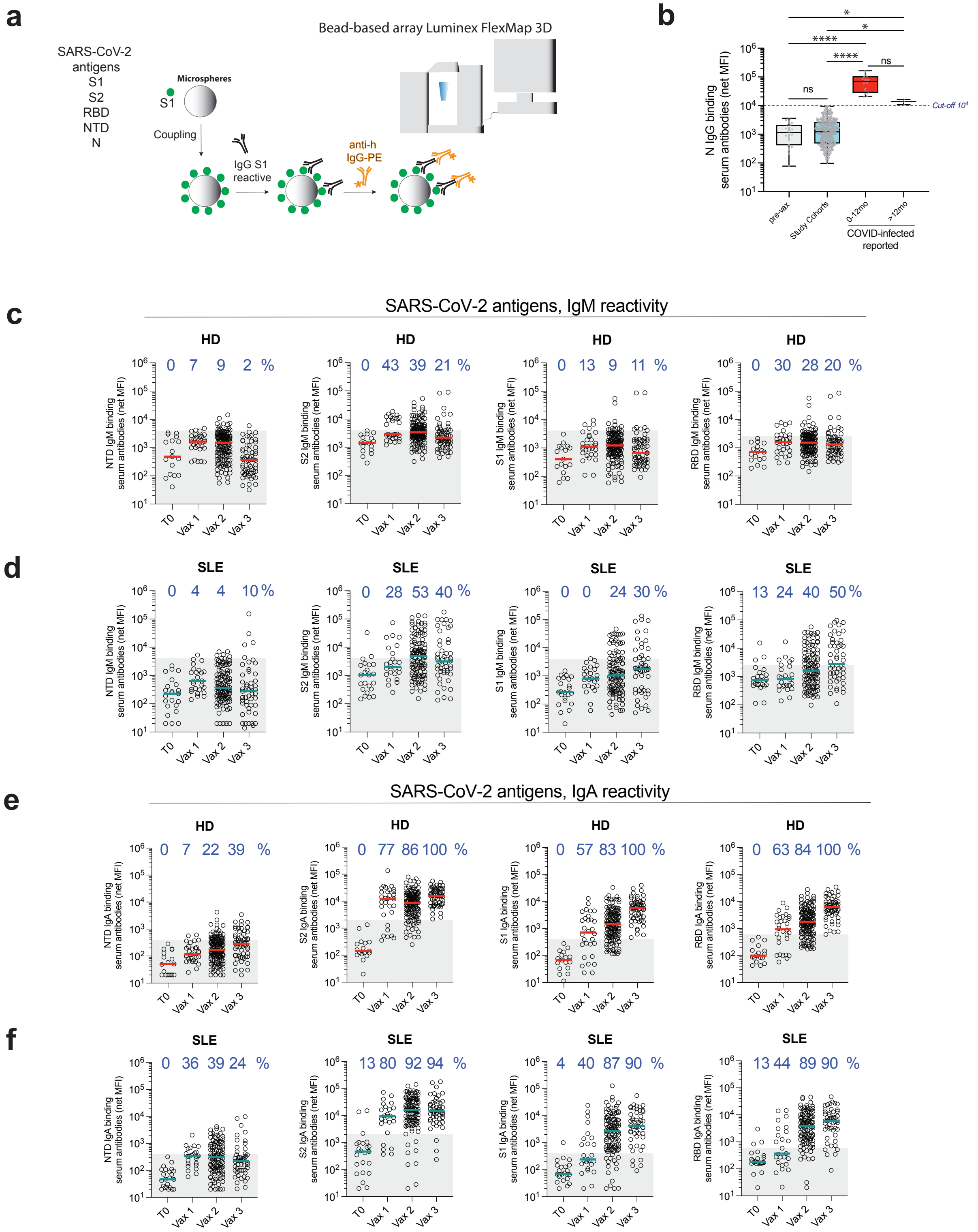

**Extended Data Fig.3 | Detection of circulating antigen specific Igs upon vaccination**

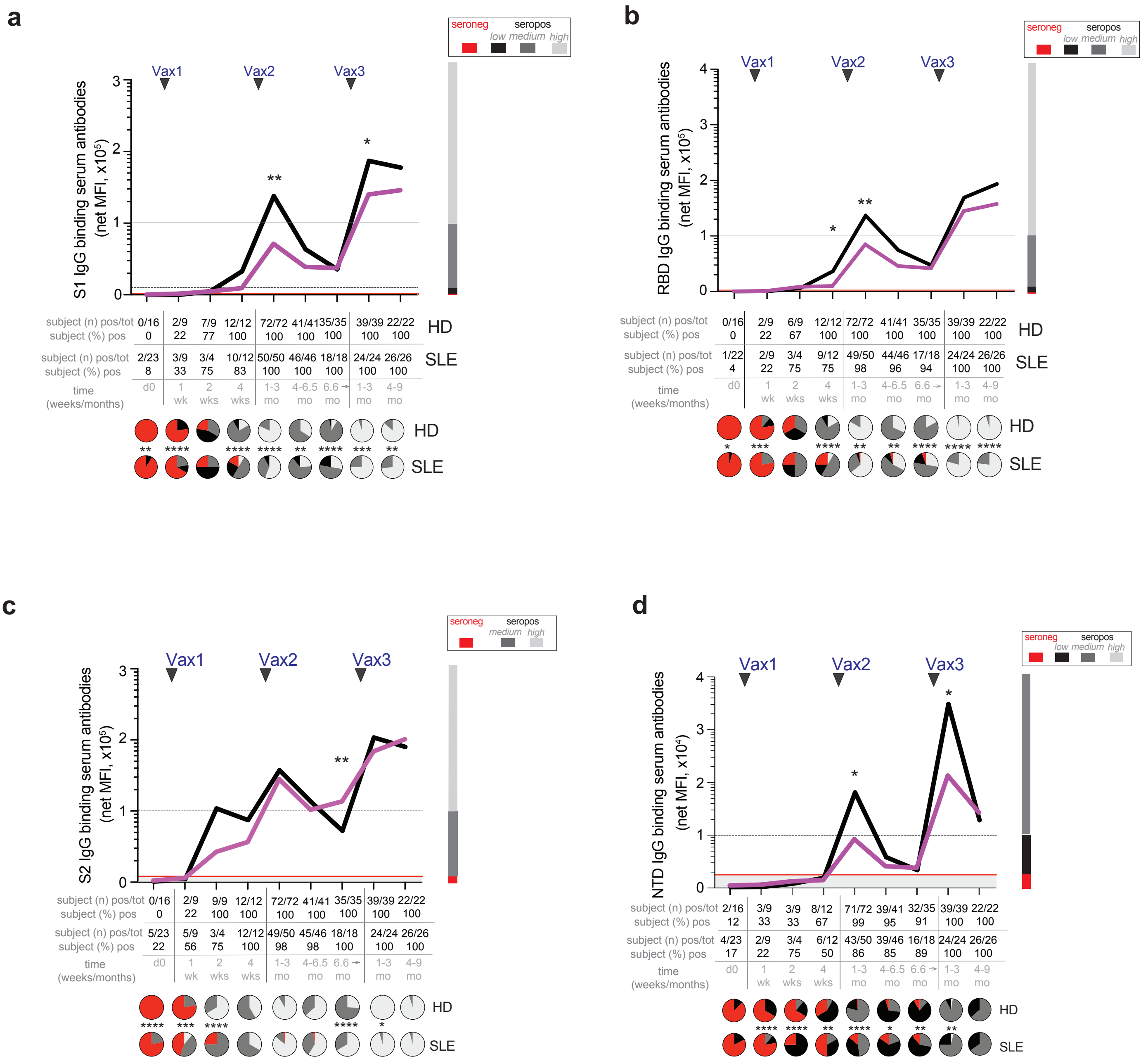

**Extended Data Fig.4 | Detection of circulating non-RBD targeting Igs upon vaccination**

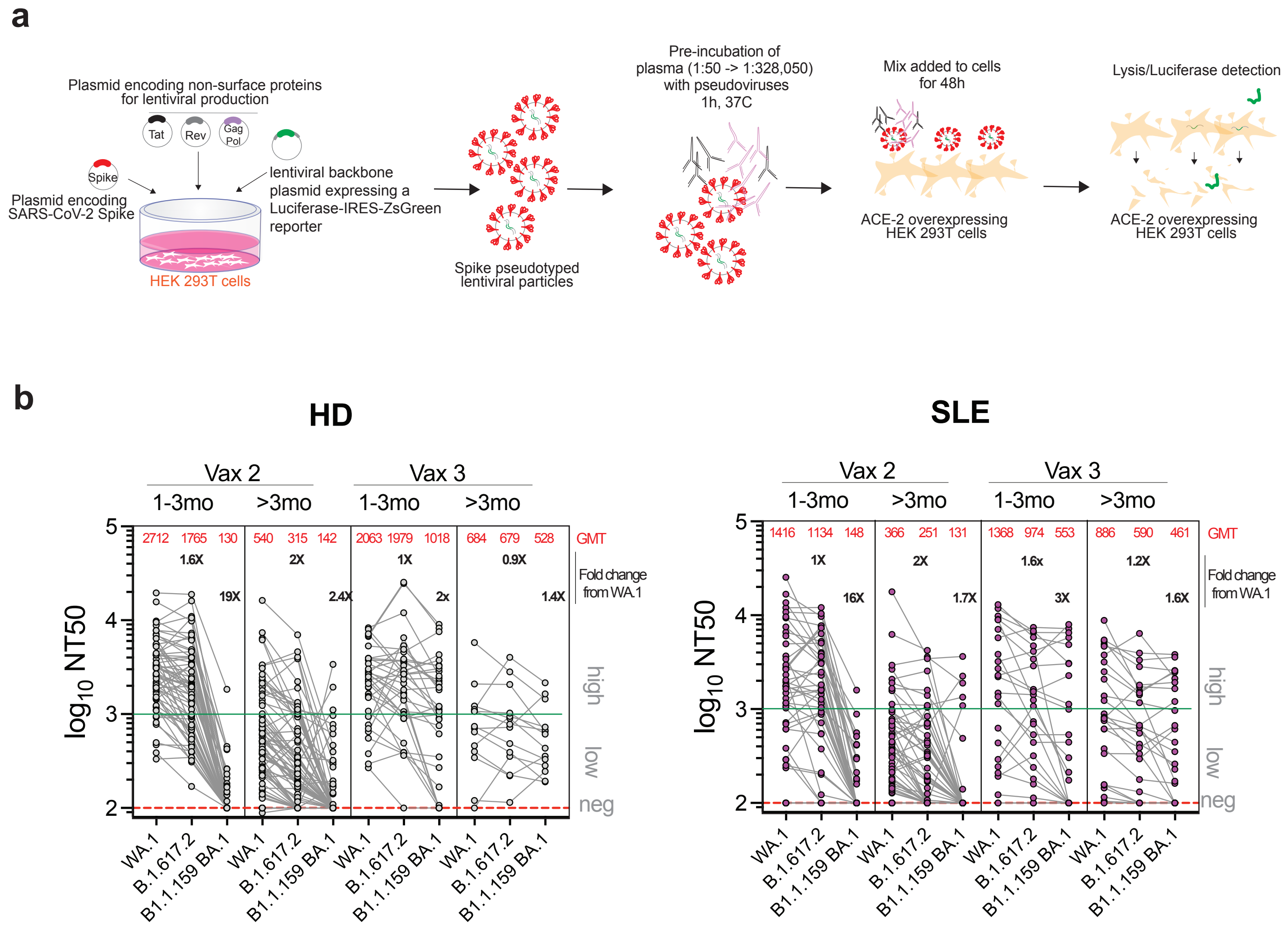

**Extended Data Fig.5 | Detection of circulating neutralizing Igs upon vaccination**

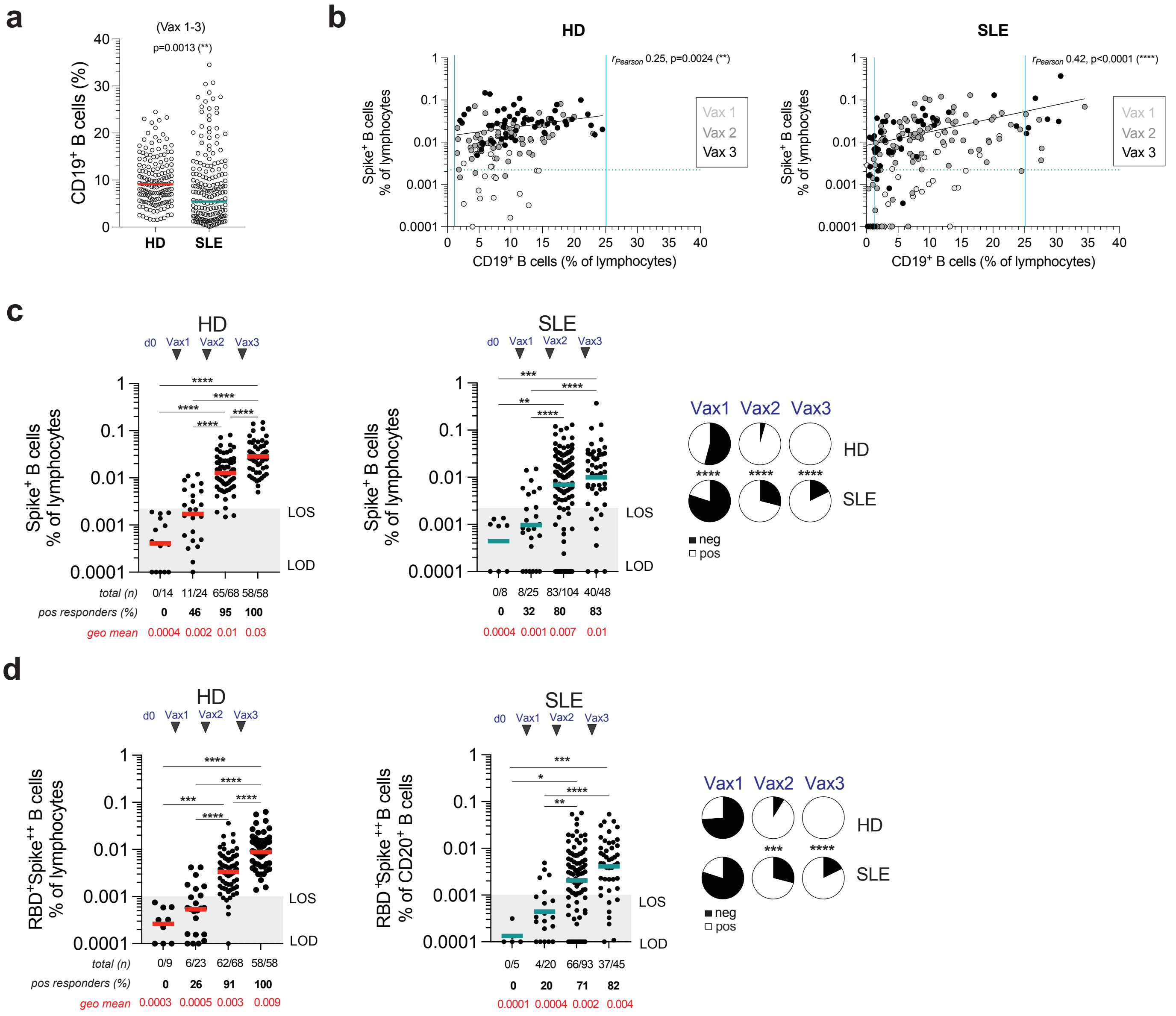

**Extended Data Fig.6 | Spike reactivity and correlation with total B cell frequencies**

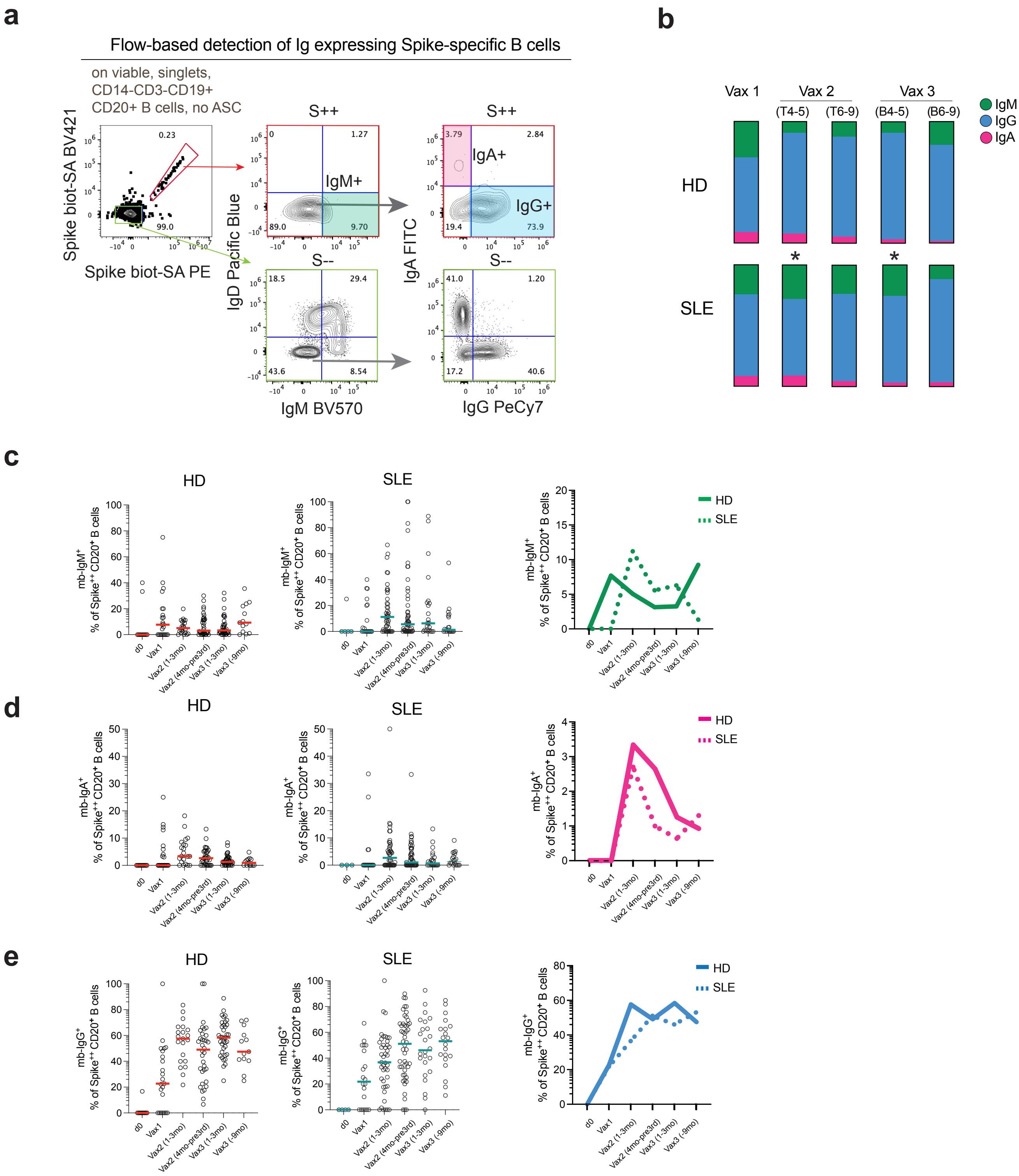

**Extended Data Fig.7 | Recall vaccine doses in SLE are characterized by a delayed expansion of csw IgG harboring B cells and increased IgM-memory reactivity towards the Spike**

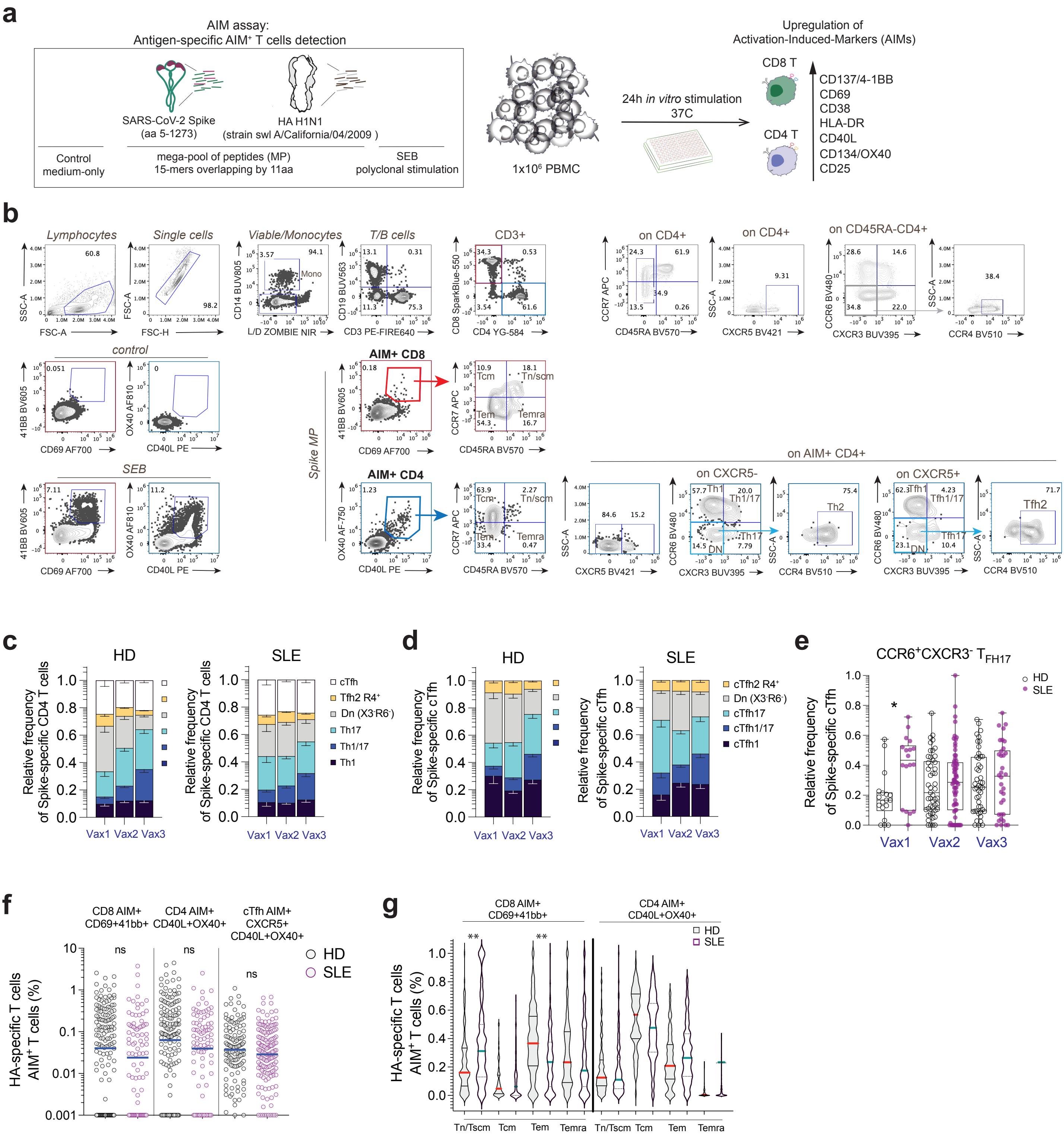

**Extended Data Fig.8 | Analysis of antigen specific T cells responses**

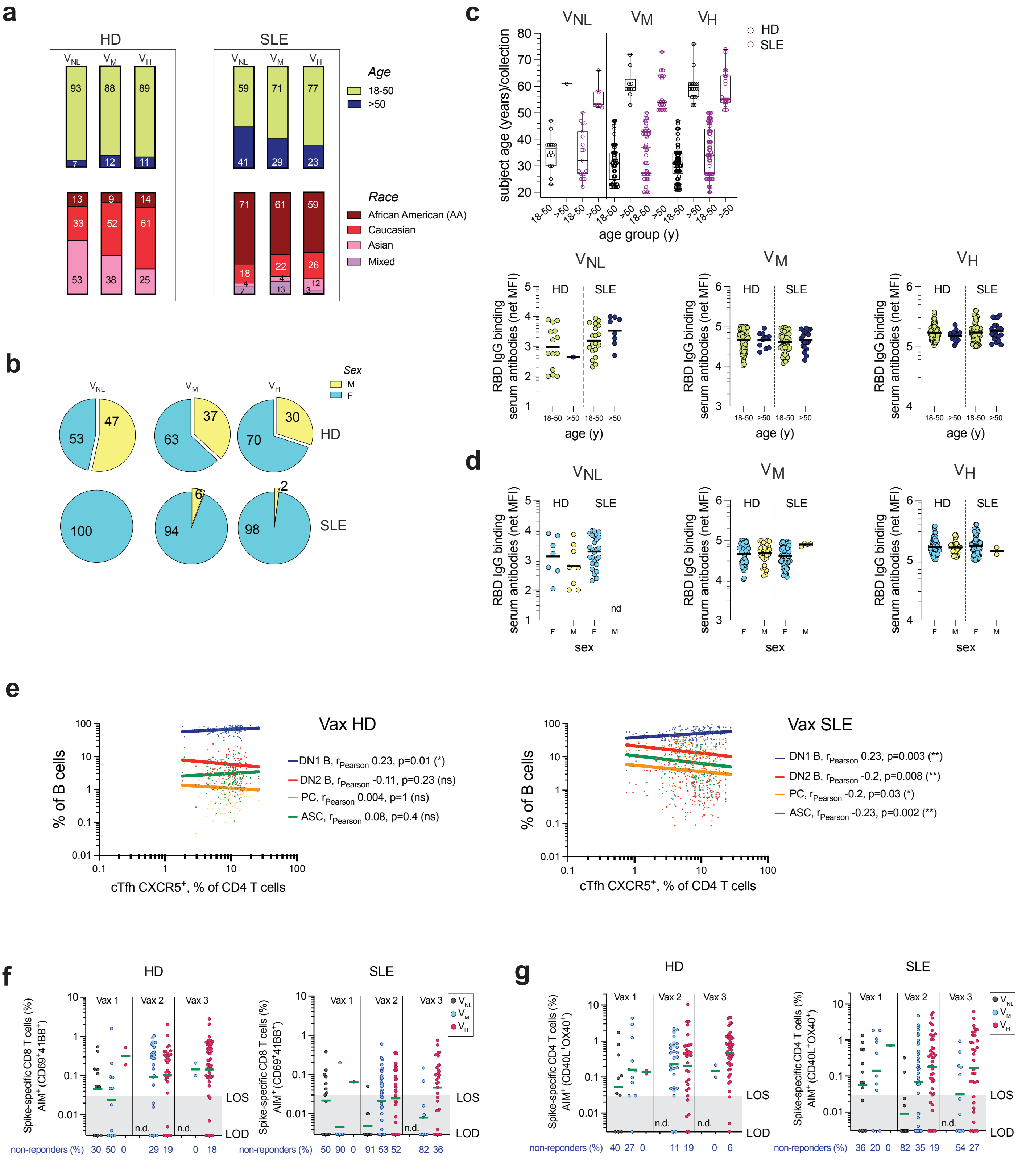

**Extended Data Fig.9 | Clinical, demographics and immunological reactivity based on vaccine-responsivness**

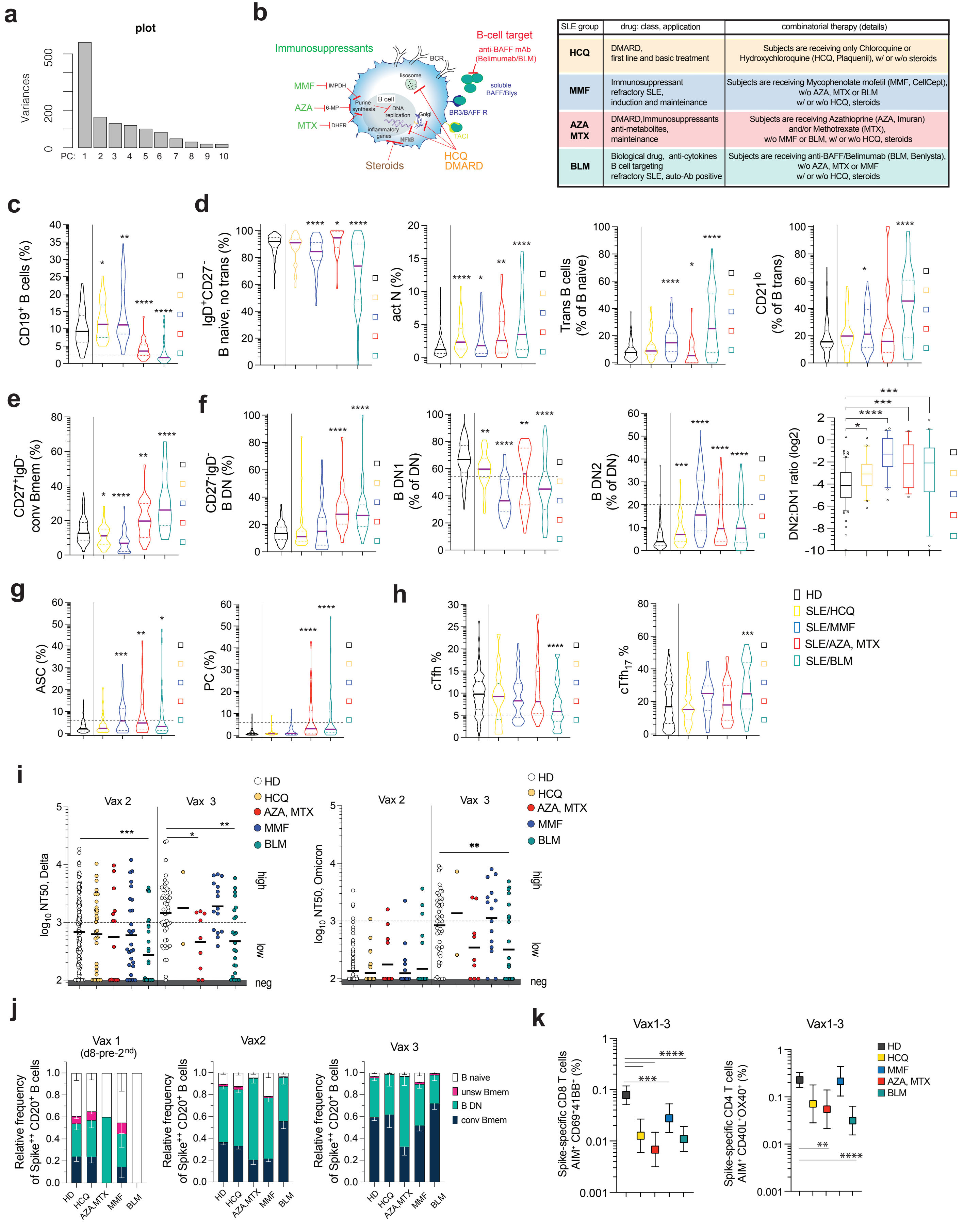

| SLE group | drug: class, application | combinatorial therapy (details) |
|---|---|---|
| HCQ | DMARD, first line and basic treatment | Subjects are receiving only Chloroquine or Hydroxychloroquine (HCQ, Plaquenil), w/ or w/o steroids |
| MMF | Immunosuppressant refractory SLE, induction and mainteinance | Subjects are receiving Mycophenolate mofetil (MMF, CellCept), w/o AZA, MTX or BLM w/ or w/o HCQ, steroids |
| AZA MTX | DMARD,Immunosuppressants anti-metabolites, mainteinance | Subjects are receiving Azathioprine (AZA, Imuran) and/or Methotrexate (MTX), w/o MMF or BLM, w/ or w/o HCQ, steroids |
| BLM | Biological drug, anti-cytokines B cell targeting refractory SLE, auto-Ab positive | Subjects are receiving anti-BAFF/Belimumab (BLM, Benlysta), w/o AZA, MTX or MMF w/ or w/o HCQ, steroids |

**Extended Data Fig.10| SLE classification based on treatments and related B and T cells immunophenotype**

Table 1. Characteristics of cohorts

| Characteristics of cohorts | | |
|---|---|---|
| | HD cohort, n= 64 | SLE cohort, n= 79 |
| Pre-COVID 19 (2016-2018) | 8 | 10 |
| Enrolled in this vaccine study | 56 | 69 |
| Gender | | |
| Female (n,%) | 38 (68) | 66 (96) |
| Male (n,%) | 18 (32) | 3 (4) |
| Age | | |
| Mean years +/- SD (range) | 38.6 +/- 14 (21-76) | 43.9 +/- 13 (20-74) |
| Race (n,%) | | |
| Black or African American | 11 (20) | 47 (68.1) |
| Asian | 12 (21) | 3 (4.35) |
| White | 33 (59) | 14 (20.3) |
| Mixed race | 0 (0) | 5 (7.25) |
| Ethnicity (n,%) | | |
| Hispanic | 1 (2) | 3 (4) |
| Non hispanic | 55 (98) | 66 (96) |
| mRNA vaccine type | | |
| Primary series (Vax1+2) | | |
| BNT162b2/BioNTech-Pfizer | 35 (66) | 43 (72) |
| mRNA-1273/Moderna | 18 (34) | 17 (28) |
| Booster (Vax3) | | |
| BNT162b2/BioNTech-Pfizer | 12 (57) | 18 (78) |
| mRNA-1273/Moderna | 9 (43) | 5 (22) |
| Interval (days) Vax3-Vax2 +/- SD | 283 +/- 53 (229-336) | 186 +/- 67 (118-253) |

Table 2. Medications of the SLE vaccinated subjects

| | |
|---|---|
| Subjects enrolled (n) | 69 |
| Current therapy | (n,%) |
| none | 1 (1) |
| Prednisone | 34 (49) |
| Prednisone<=20 mg/day | 34 (100) |
| Prednisone >=20 mg/day | 0 (0) |
| Hydroxychloroquine | 59 (85) |
| 200 mg | 30 (51) |
| 300-400 mg | 28 (49) |
| Immunosuppresive drugs | |
| Azathioprine (AZA) | 12 (17) |
| 50-100 mg | 8 (67) |
| 150 mg | 4 (33) |
| Methotrexate (MTX) | 2 (3) |
| 15 mg/week | 2 (100) |
| Mycophenolate Mofetil (MMF) | 20 (29) |
| 500 mg | 2 (10) |
| 1000-1800 mg | 7 (35) |
| 2000-3000 mg | 11 (55) |
| Biologic therapy | |
| Belimumab | 22 (32) |
| subQ injection (weekly) | 15 (68) |
| IV injection (monthly) | 7 (32) |
